# Visuomotor brain network activation and functional connectivity among individuals with autism spectrum disorder

**DOI:** 10.1101/2021.03.31.21254423

**Authors:** Rebecca J. Lepping, Walker S. McKinney, Grant C. Magnon, Sarah K. Keedy, Zheng Wang, Stephen A. Coombes, David E. Vaillancourt, John A. Sweeney, Matthew W. Mosconi

**Author notes:** Submitting and corresponding author information: Matthew W. Mosconi, Ph.D., Dole Human Development Center, 1000 Sunnyside Ave., Office 2018, Lawrence, KS 66045.

## Abstract

Sensorimotor abnormalities are common in autism spectrum disorder (ASD) and predictive of functional outcomes, though their neural underpinnings remain poorly understood. Using functional magnetic resonance imaging (fMRI), we examined both brain activation and functional connectivity during visuomotor behavior in 27 individuals with ASD and 30 typically developing (TD) controls (ages 9-35 years). Participants maintained a constant grip force while receiving visual feedback at three different visual gain levels. Relative to controls, ASD participants showed increased force variability, especially at high gain, and reduced entropy. Brain activation was greater in individuals with ASD than controls in supplementary motor area, bilateral superior parietal lobules, and left middle frontal gyrus at high gain. During motor action, functional connectivity was reduced between parietal-premotor and parietal-putamen in individuals with ASD compared to controls. Individuals with ASD also showed greater age-associated increases in functional connectivity between cerebellum and visual, motor, and prefrontal cortical areas relative to controls. These results indicate that visuomotor deficits in ASD are associated with atypical activation and functional connectivity of posterior parietal, premotor, and striatal circuits involved in translating sensory feedback information into precision motor behaviors, and that functional connectivity of cerebellar-cortical sensorimotor and non-sensorimotor networks show delayed maturation.

**HIGHLIGHTS:**

- Individuals with ASD show impaired precision manual force control, especially when visual feedback is magnified
- Visuomotor deficits in ASD are associated with increased activity in frontal and parietal cortex and reduced parietal-frontal and parietal-putamen functional connectivity
- Visuomotor-dependent functional connectivity of cerebellum with visual, motor, and prefrontal cortices shows atypical age-associated trajectories in ASD

## 1. INTRODUCTION

Autism spectrum disorder (ASD) affects the development of multiple cognitive and behavioral abilities. The diverse range of clinical issues associated with ASD hinders progress identifying neurodevelopmental mechanisms. Sensorimotor behaviors represent a promising target for advancing knowledge of neurodevelopmental mechanisms of ASD because they are frequently disrupted (Coll et al. 2020), predictive of worse outcomes (Nebel et al. 2016; Marrus et al. 2018), familial (Mosconi et al. 2010; Schmitt et al. 2019), and supported by well understood and highly specialized brain networks. Quantitative studies of sensorimotor behavior and brain function in ASD may advance a more mechanistic understanding of the disorder.

Individuals with ASD show a range of sensorimotor abnormalities including repetitive motor mannerisms (American Psychiatric Association 2013), less precise saccadic eye movements (Takarae et al. 2004; Johnson et al. 2013; Schmitt et al. 2014), increased postural sway (Wang et al. 2016; Bojanek et al. 2020), and dyspraxia (MacNeil and Mostofsky 2012). Multiple studies converge to suggest that individuals with ASD show reduced ability to integrate visual or multi-sensory feedback during motor behavior (Haswell et al. 2009; Sharer et al. 2015). Consistent with this hypothesis, we have found that individuals with ASD show greater motor variability and reduced entropy during visually guided precision gripping implicating deficient visual feedback control of motor behavior (Mosconi et al. 2015; Wang et al. 2016; Neely et al. 2019).

During visually guided motor behavior (*i.e.,* visuomotor behavior), visual feedback is processed in visual and posterior parietal cortices, including superior (SPL) and inferior parietal lobules (IPL), which project to premotor and primary motor cortices (M1; Caminiti et al., 1996; Vaillancourt et al., 2006b). Visuomotor behavior is supported by a subcortical circuit in which feedback information is relayed from posterior parietal cortex to cerebellum where error information is translated into reactive motor commands to M1 (Stein and Glickstein 1992; Glickstein 2000; Glover et al. 2012). Posterior cerebellum and basal ganglia, including putamen and caudate, are involved in modulating motor output timing and amplitude (Prodoehl et al. 2008; Spraker et al. 2012). The function of these networks is consolidated throughout childhood and early adulthood as evidenced by strengthening of long-distance cerebellar-cortical pathways and weakening of local functional circuits (Amemiya et al. 2019). Developmental studies of visuomotor network function in ASD identified increases in cerebellar-cortical functional connectivity in childhood (Stoodley et al. 2017) that become more severe during adolescence and adulthood (Holiga et al. 2019). Despite evidence that task-based functional magnetic resonance imaging (fMRI) approaches to assessing functional connectivity provide more robust predictors of behavioral trait dimensions than resting state (Greene et al. 2018), few studies have assessed task-dependent functional connectivity of visuomotor networks in ASD or their variance across childhood and adulthood.

Several studies have assessed visuomotor network activation in ASD. Takarae and colleagues (2007) reported reduced frontal and parietal eye field and cerebellar activation during saccades in adults with ASD relative to typically developing (TD) controls. Individuals with ASD also showed greater activity in frontal, striatal and cerebellar regions suggesting increased involvement of cognitive control networks. Finger tapping studies have also documented aberrant frontal, parietal and cerebellar activity in ASD (Muller et al. 2001; Allen and Courchesne 2003; Muller et al. 2003; Allen et al. 2004; Mostofsky et al. 2009). In the one known study to assess functional connectivity during motor behavior, Mostofsky and colleagues (2009) documented reduced cerebellar-thalamo-cortical functional connectivity alongside increased supplementary motor area activation and reduced cerebellar activation in ASD during finger tapping. While finger-tapping studies have been important for identifying gross motor impairment, studies of fine motor control, such as during precision gripping, allow for interrogation of the visuomotor system in a way that more accurately represents functional motor behaviors. We previously have described fMRI task activation increases in ventral premotor cortex and cerebellum during a precision gripping task in ASD (Unruh et al. 2019). While the majority of motor behaviors are guided by visual feedback, and visuomotor behaviors are impaired in ASD, no known fMRI studies have examined brain network function and connectivity in ASD during visually guided precision motor behavior.

Using fMRI, we assessed visuomotor network activation and functional connectivity during precision gripping. We predicted that, relative to TD controls, individuals with ASD would show increased grip force variability and reduced force entropy. Based on prior behavioral findings that visuomotor impairments in ASD are more severe at high visual feedback gain (Mosconi et al. 2015), we tested behavior and brain function across three visual gain levels. We predicted that increases in force variability and reductions in entropy would be more severe when visual feedback was amplified indicating deficits processing increased sensory information. Consistent with the overarching hypothesis that visuomotor deficits in ASD reflect alterations in dynamically adjusting motor behavior in response to sensory feedback, we predicted individuals with ASD would show reduced activation of visual cortex, posterior parietal cortex, and cerebellum during visuomotor control relative to TD controls. We also expected reduced parietal-cerebellar, parietal-striatal, and parietal-motor cortex functional connectivity in ASD during visuomotor behavior. Individuals were studied across a wide age range (9-35 years) so that age-associated differences in visuomotor behavior and brain network function could be examined. Consistent with prior resting state fMRI studies (Padmanabhan et al. 2013), we predicted visuomotor brain dysfunctions relative to TD controls would be more severe at older ages.

## 2. MATERIALS AND METHODS

### 2.1 Participants

Twenty-seven participants with ASD and 30 TD controls were recruited from the community, and group-matched on age (range: 9-35 years), nonverbal IQ, and handedness (Table 1). IQ was assessed using the Wechsler Abbreviated Scale of Intelligence (WASI; Wechsler 1999). ASD diagnoses were confirmed using the Autism Diagnostic Inventory-Revised (ADI-R; Lord et al. 1994), the Autism Diagnostic Observation Schedule (ADOS-2; Lord et al. 1989) and expert clinical opinion using DSM-IV-TR criteria (American Psychiatric Association 2000). Participants with ASD were excluded if they had a known genetic or metabolic disorder associated with ASD (*e.g.*, fragile X syndrome, tuberous sclerosis). TD control participants were excluded if they scored higher than eight on the Social Communication Questionnaire (SCQ; Berument et al. 1999), or reported current or past psychiatric or neurological disorders, family history of ASD in first-, second- or third-degree relatives, or first-degree relative with a developmental or learning disorder, psychosis, or obsessive-compulsive disorder. No participants were taking medications known to affect motor control, including antipsychotics, stimulants, or anticonvulsants (Reilly et al. 2005), nor had a history of head injury, birth injury, or seizure disorder. Participants’ uncorrected far visual acuity was at least 20/40. Written informed consent was obtained for adult participants and parental consent was obtained for participants younger than 18 years. Minors provided written assent. Study procedures were reviewed and approved by the Institutional Review Board of the University of Illinois – Chicago and abided by the Code of Ethics of the World Medical Association (Declaration of Helsinki).

**Table 1.**
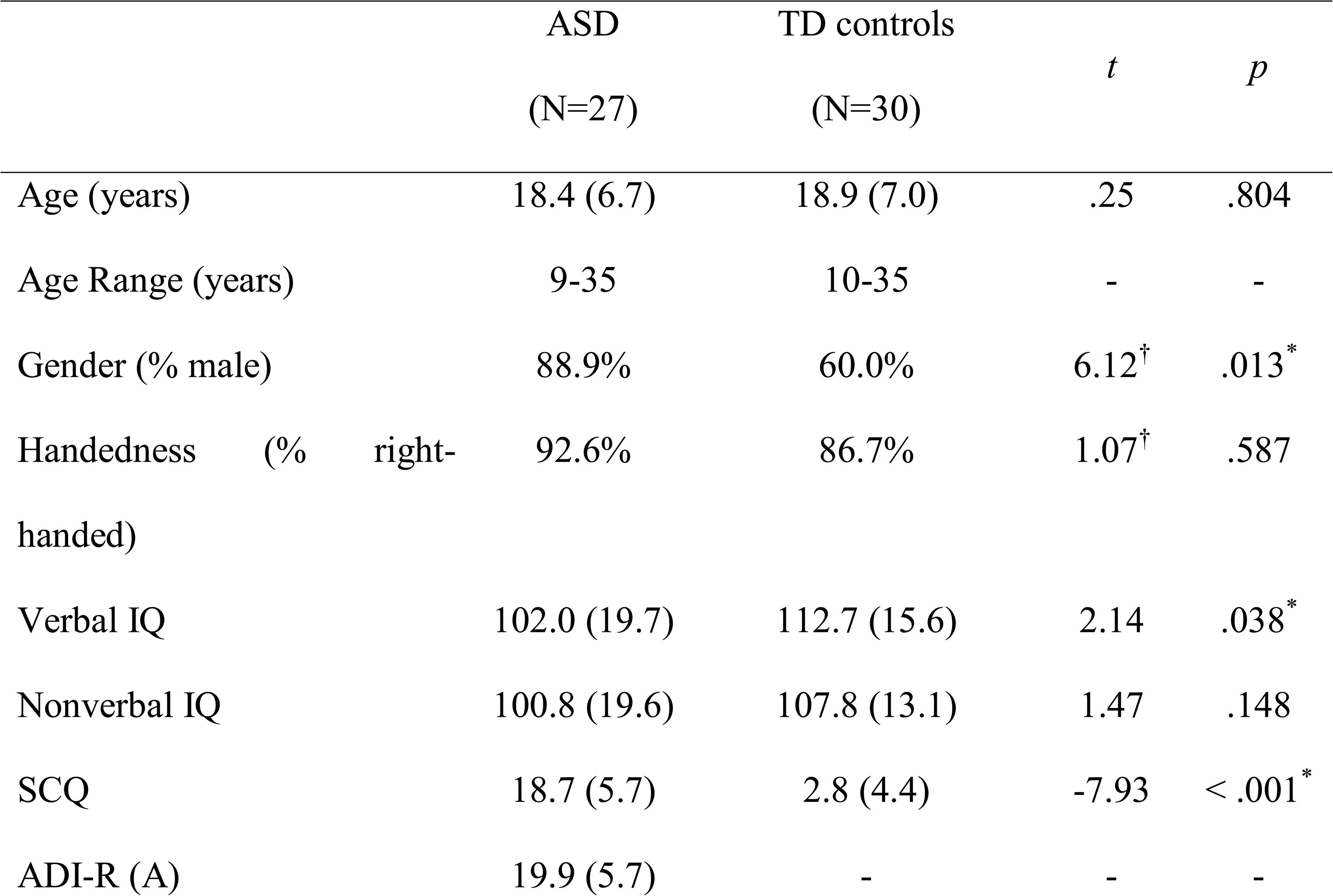
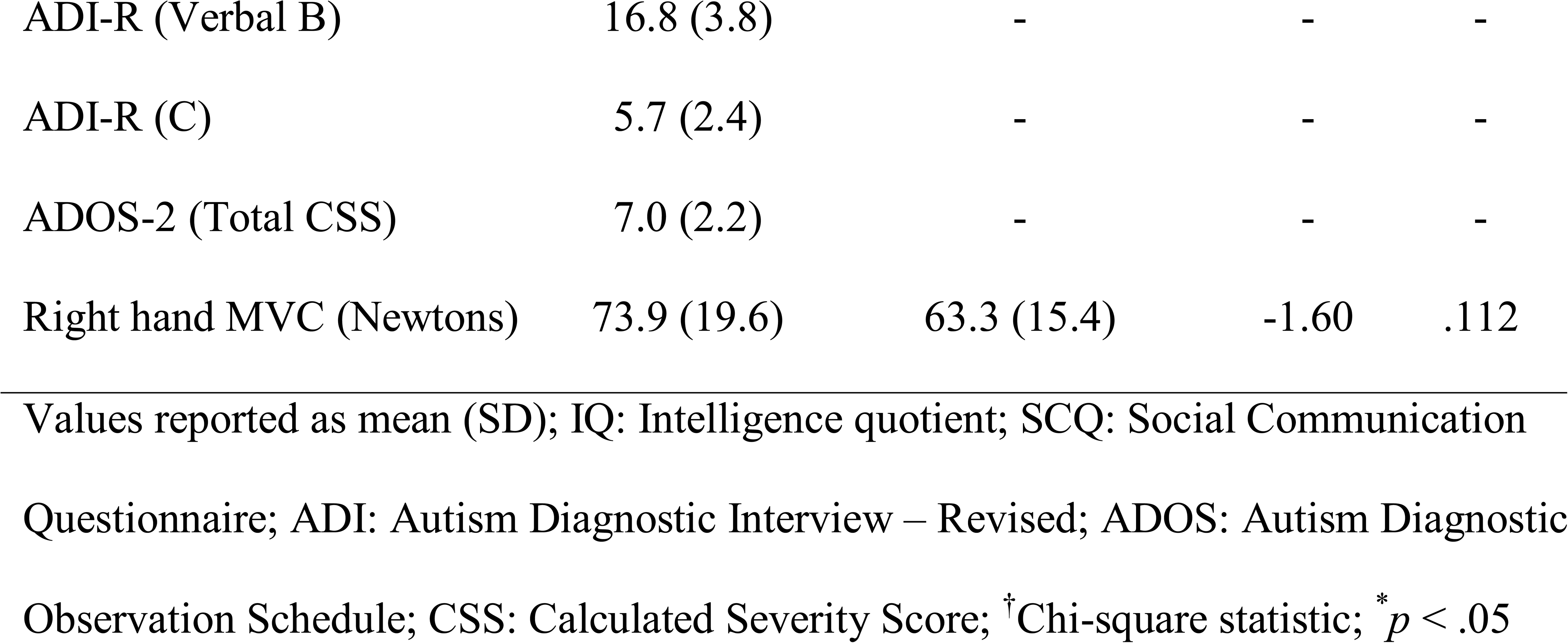
Demographic characteristics for individuals with ASD and TD controls

|  | ASD<br>(N=27) | TD controls<br>(N=30) | *t* | *p* |
| --- | --- | --- | --- | --- |
| Age (years) | 18.4 (6.7) | 18.9 (7.0) | .25 | .804 |
| Age Range (years) | 9-35 | 10-35 | - | - |
| Gender (% male) | 88.9% | 60.0% | 6.12<sup>†</sup> | .013<sup>*</sup> |
| Handedness (% right-handed) | 92.6% | 86.7% | 1.07<sup>†</sup> | .587 |
| Verbal IQ | 102.0 (19.7) | 112.7 (15.6) | 2.14 | .038<sup>*</sup> |
| Nonverbal IQ | 100.8 (19.6) | 107.8 (13.1) | 1.47 | .148 |
| SCQ | 18.7 (5.7) | 2.8 (4.4) | -7.93 | < .001<sup>*</sup> |
| ADI-R (A) | 19.9 (5.7) | - | - | - |
| ADI-R (Verbal B) | 16.8 (3.8) | - | - | - |
| ADI-R (C) | 5.7 (2.4) | - | - | - |
| ADOS-2 (Total CSS) | 7.0 (2.2) | - | - | - |
| Right hand MVC (Newtons) | 73.9 (19.6) | 63.3 (15.4) | -1.60 | .112 |

Values reported as mean (SD); IQ: Intelligence quotient; SCQ: Social Communication
Questionnaire; ADI: Autism Diagnostic Interview – Revised; ADOS: Autism Diagnostic
Observation Schedule; CSS: Calculated Severity Score; <sup>†</sup>Chi-square statistic; \* $p < .05$

### 2.2 Data Acquisition

#### 2.2.1 Visuomotor Behavior Data Acquisition

Participants used their right thumb and index finger to exert opposing forces on a custom fiber-optic force transducer (Figure 1A; Neuroimaging Solutions), constructed from rigid, nonmetallic material to ensure safety and consistent linearity, sensitivity, and accuracy inside the MR environment. The force signal was transmitted from the transducer via fiber-optic cable to a si425 Optical Sensing Interrogator (Micron Optics), which digitized the force data at 125 Hz. Customized software written in LabVIEW (National Instruments, Austin, TX) converted analog force data to Newtons (N) at a resolution of 0.025 N. Output from the force transducer was presented to the participant using a visual display through a projection system placed on the head coil and a mirror located 35 cm from the participant’s eyes inside the MR environment (resolution: 640 × 480 pixels; refresh rate: 60 Hz).

**Fig 1.**
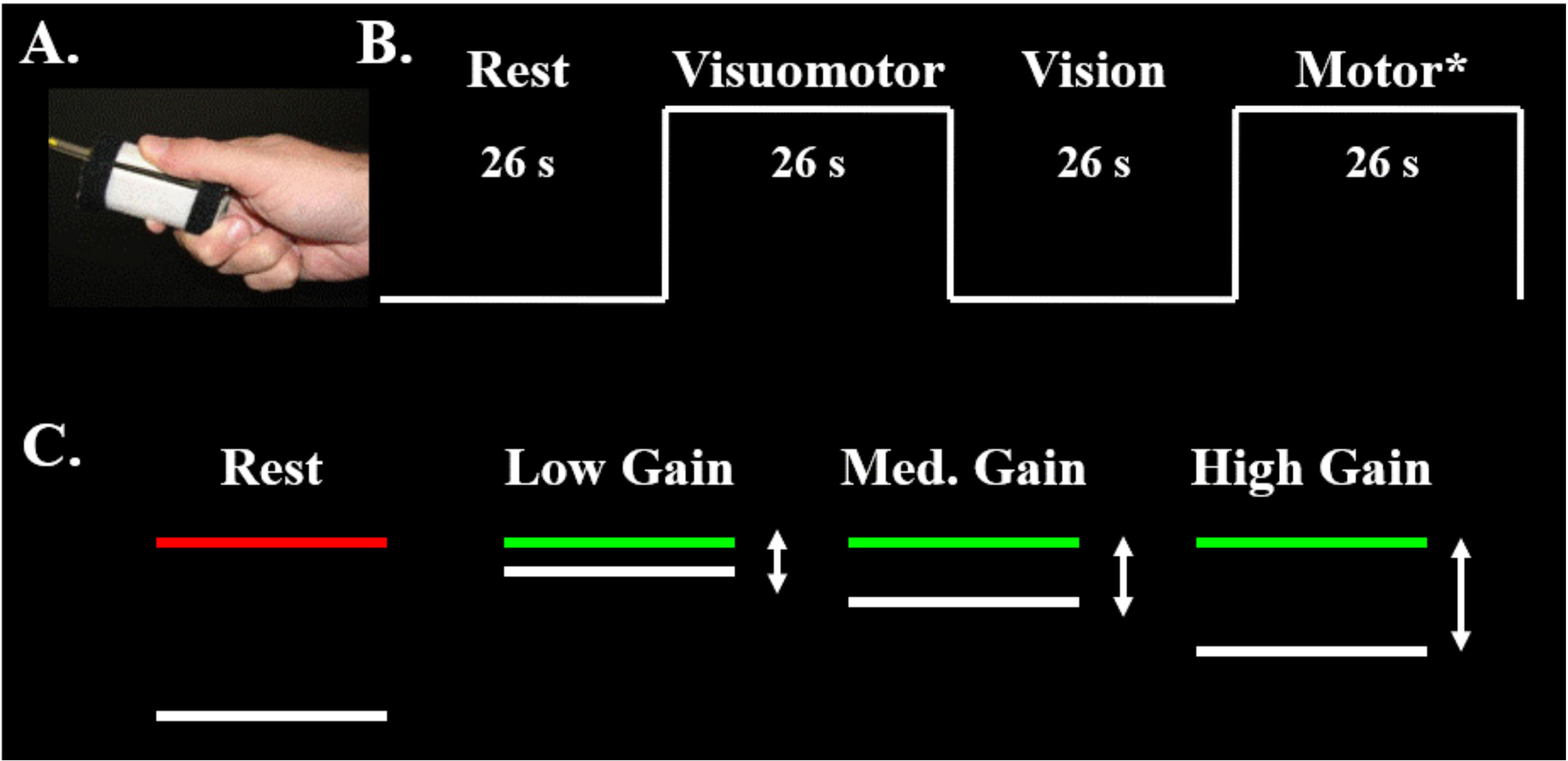
A) Grip configuration and force transducer. B) Schematic representation of task condition blocks. Blocks are 26 seconds in duration. Each condition was repeated three times at each gain level, ending each run with a rest block. C) Schematic representation of the visuomotor task. The red target bar turned green to cue the beginning of each trial. The white force bar moved upwards with increased force. The force bar traveled a greater distance per change in force for higher relative to lower gains. *results from the *motor only* condition in which no visual feedback was provided are not reported due to participants’ difficulties remembering to press.

#### 2.2.2 MRI Data Acquisition

MR scans were performed using a 3 Tesla scanner (General Electric Medical Systems, Milwaukee, WI) with a quadrature head coil. Participants lay supine and their heads were stabilized using adjustable padding. Functional images were acquired using a T2* single shot gradient-echo echo-planar pulse sequence: repetition time (TR) = 2000 ms, echo time (TE) = 25 ms, flip angle = 90°, in-plane resolution = 3 × 3 mm, 64 × 64 acquisition matrix, field of view (FOV) = 200 mm, 33 axial slices, 3 mm thickness with 1 mm gap. An anatomical scan was acquired using a T1-weighted 3D inversion recovery fast spoiled gradient recalled pulse (SPGR) sequence: TR = 25 ms, TE = 3 ms, flip angle = 90°, in-plane resolution = 0.9 × 0.9 mm, 256 × 192 acquisition matrix, FOV = 24 × 24, 120 axial slices, 1.5 mm thickness with no gap. Both sequences covered the entire brain.

### 2.3 Experimental Design

Participants completed a 30-minute pre-fMRI training session completing at least one run of the fMRI task. The training session ensured participants understood the instructions and were able to complete the task without verbal cues, and also helped eliminate the transitory period of motor learning (Coombes et al. 2010). Each participant’s maximum voluntary contraction (MVC) was calculated before beginning the fMRI experiment. Participants produced their maximum force against a strain-gauge dynamometer over three trials (Sammons Preston, Rolyan, Bolingbrook, IL). MVC was estimated as the mean maximum value (Vaillancourt et al. 2006b; Mosconi et al. 2015).

Three fMRI task runs were administered. During each run, participants completed four task conditions with their right hand in the following order: 1) rest, 2) visuomotor, 3) vision, and 4) motor only. Each condition block was 26 seconds long. The condition series was administered three times, with an additional *rest* block at the end of each run (*e.g., R-VM-V-M-R-VM-V-M-R-VM-V-M-R*; Figure 1B). The duration of each run was 5:38. Each of the three runs followed the same sequence, but the visual gain of feedback was varied. The target force was fixed at 15% of the MVC. During the *rest* condition, participants viewed a horizontal white force bar and a parallel red target bar (Figure 1B-C). They were instructed to watch the static bars, and ensure they did not press the transducer. During the *visuomotor* condition, the red target bar turned green at trial onset, and the white force bar moved upwards with increased force and downward with decreased force. Participants were instructed to press when the red target bar turned green, and to keep pressing so that the white force bar stayed as steady as possible matching the green target bar. *Vision only* trials were administered to assess blood oxygenation level dependent (BOLD) responses to visual motion without force production. Participants viewed the green target bar and the moving force bar, but did not produce force. The force bar oscillated around the target bar, and participants were instructed to watch the screen. The oscillation frequency of the force bar was a 1-Hz sine wave with a small amount of white noise. The amplitude of oscillation matched the visual gain of the run being administered. The *motor only* condition was administered to assess BOLD responses to motor behavior without visual feedback. During this condition, the target bar remained green, but the force bar disappeared after 1.5 seconds. Participants were instructed to continue pressing with the same level of force used to reach the target. Due to participants’ difficulties remembering to press during the motor only condition, these data are not reported here.

Visual gain was manipulated by changing the visual angle. Visual angle (α) was varied by altering the height of force fluctuations (H_1_) on the video display, relative to the distance between the participant’s eyes and screen (D) using the following formula (Vaillancourt et al. 2006a):

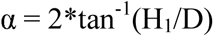

Based on prior studies, we assumed that participants produced force at 6 N with a standard deviation (SD) of 0.3 N (Slifkin and Newell 1999; Mosconi et al. 2015). SD multiplied by six approximated the full range of estimated variance. H_1_ was varied to approximate three visual angles: 0.018° (low gain), 0.192° (medium gain) and 2.023° (high gain) to obtain values above and below one degree (Vaillancourt et al. 2006b). The order of administration of gain levels was counterbalanced across participants.

### 2.4 Data Processing

#### 2.4.1 Visuomotor Behavior Data Processing

To examine visuomotor performance, the behavior time series data were processed using a custom LabVIEW program (National Instruments, Austin, TX) (Elliott et al. 2007) and MATLAB (The MathWorks Inc., Natick, MA). Time series data for each fMRI task run were digitally filtered using a fourth-order Butterworth filter with a 30 Hz low-pass cutoff. To examine sustained force output, the first two seconds and last second of the force trace were excluded for each 26 second visuomotor block. The trace was then linearly detrended to account for changes in mean force over the course of the trial. Mean force was calculated as the average force output of the time series as a measure of individuals’ ability to complete the task. The within-trial SD of the force time series was calculated to examine the amplitude of performance variability. To examine the time-dependent structure of the data, approximate entropy (ApEn) was calculated for each trial (Pincus and Goldberger 1994; Vaillancourt et al. 2001). ApEn reflects the predictability of future values based on previous values. For example, a sine wave has accurate short- and long-term predictability and corresponds to an ApEn near zero. Increases in signal complexity, reflective of the independence of each force value, returns an ApEn near two (*e.g.* white noise). The same algorithm and parameter settings (*m* = 2; *r* = 0.2*SD of the signal) were applied as in our previous work (Slifkin et al. 2000; Mosconi et al. 2015; McKinney et al. 2019; Unruh et al. 2019).

#### 2.4.2 fMRI Data Processing

Imaging data were processed using the Analysis of Functional Neuroimages software (AFNI; https://afni.nimh.nih.gov; Cox 1996). MR data were rejected for head movement artifact and failure to comply with task instructions for seven participants’ low gain condition (ASD:4, TD:3), four participants’ medium gain condition (ASD:3, TD:1), and six participants’ high gain condition (ASD:4, TD:2). Participants were included in group analyses if they successfully completed at least one gain condition. Anatomical images were skull-stripped and nonlinearly warped to Montreal Neurologic Institute (MNI) standard space (Fonov et al. 2011). Functional preprocessing steps followed our previously reported work (Unruh et al. 2019; McKinney et al. 2020). Slice-timing correction was applied. Consecutive volumes with > 0.5 mm framewise displacement were censored. Motion censoring data are in Supplementary Table 1. The percent of TRs censored (gain effect: *F*_(2,95.21)_ = 0.74, *p* = .48) and average motion per TR after censoring (gain effect: *F*_(2,95.64)_ = 2.48, *p* = .09) were similar across gain levels and groups (percent of TRs censored group effect: *F*_(1,51.78)_ = 2.05, *p* = .16; average motion per TR group effect: *F*_(1,51.91)_ = 1.48, *p* = .23). Group differences did not vary as a function of gain for either percent of TRs censored (group × gain interaction: *F*_(2,95.21)_ = 0.31, *p* = .73) nor average motion per TR (group × gain interaction: *F*_(2,95.63)_ = 0.74, *p* = .48).

Remaining functional volumes were rigidly aligned with anatomical data referenced to the minimum outlier volume and warped into standard space. Volumes were spatially smoothed to a 6 mm full-width half-maximum Gaussian kernel and scaled to the mean voxel time-series value of 100. Functional data were regressed using a block function with six motion parameters (X, Y, Z, roll, pitch, yaw) included as nuisance regressor terms. Regression outcomes represent the percent signal change (*β*) of each contrast of interest (visuomotor – rest; vision – rest; visuomotor – vision) and associated *t*-statistics.

Separate psychophysiological interaction (PPI), or task-dependent functional connectivity analyses (McLaren et al. 2012; Cisler et al. 2014) were conducted for eight hypothesis-driven seed regions of interest (ROIs) identified from previous precision gripping studies (Vaillancourt et al. 2006b; Spraker et al. 2012), including bilateral IPL, SPL, cerebellar Crus I, and cerebellar lobules V/VI (Supplementary Figure 1). Cortical ROIs were obtained using Brainnetome, a cortical atlas parcellated from resting-state functional connectivity data of 40 healthy, right-handed adults (Fan et al. 2016). Cerebellar ROIs were obtained using the spatially unbiased infratentorial template (SUIT) cerebellar atlas (Diedrichsen 2006), created by averaging high-resolution cerebellar/brainstem scans of 20 adults. The average timeseries of each seed ROI was calculated. A canonical hemodynamic response function (HRF) was calculated for visuomotor, vision, and rest blocks, as were interactions between the ROI timeseries and visuomotor, vision, and rest functions (*i.e.*, PPI regressors). For each seed region, the seed timeseries, the three HRFs, the three PPI regressors, and 12 motion regressors were included in regression models using AFNI’s *3dDeconvolve* program. Visuomotor-dependent functional connectivity for PPI contrasts of interest (visuomotor – rest) was calculated using AFNI’s *3dcalc* program.

### 2.5 Statistical Analysis

Age, IQ, and MVC were compared between groups (ASD vs. TD controls) using two-sided independent samples *t*-tests. Handedness and sex were compared between groups using chi-square tests. Non-normally distributed behavioral outcomes (force SD, ApEn) were log-transformed. A series of linear mixed effects models were performed to examine group differences in behavioral outcomes across visual gain levels (low, medium, and high). Mixed effects models were used to estimate missing data and model within subject variation in outcome measures.

Group differences in brain activation and visuomotor-dependent functional connectivity were identified with linear mixed effects modeling using AFNI’s *3dLME* program (Chen et al. 2013). Separate 2 (group) × 3 (gain) models with group × gain and group × age interaction terms were examined for visuomotor – rest, vision – rest, and visuomotor – vision contrasts. Sex was included in each model as a covariate of no interest. A group-level mask (*3dmask_tool*) was applied to all *3dLME* output to include voxels present in at least 50% of the data sets. Auto-correlation function (ACF) estimates derived from residual data were averaged across the sample and entered into AFNI’s *3dClustSim* program to estimate family-wise error correction at α < .05. Based on this, we report significant clusters with at least 57 contiguous voxels (1539 mm^3^) at voxel-wise *p* < .005 for group contrasts and group interactions. To better differentiate large clusters of activation observed in main effect tests of visual feedback gain, a more stringent voxel-wise threshold of *p* < .001 and 23 contiguous voxels (621 mm^3^) was used to achieve corrected α < .05.

Linear mixed effects models were conducted to examine relationships between brain outcomes and behavioral and clinical variables. Maximum BOLD signal for individual participants for each significant cluster in group-contrast analyses was extracted using the clusters as masks. Comparison variables included force SD and ApEn (both groups); and clinical measures (ASD only), including scores for the ADOS (overall calculated severity score) and ADI (diagnostic algorithm scores for each subscale). Due to the exploratory nature of these analyses, we report as significant all relationships with uncorrected *p* < .05.

## 3. RESULTS

### 3.1 Visuomotor Behavior Performance

Individuals with ASD and controls showed similar MVCs (Table 1; ASD: range: 40-110 N; TD: range: 34-88 N). Mean force was similar across gain levels (gain main effect: *F*_(2,104.31)_ = 0.06, *p* = .94) and groups (Figure 2A; group main effect: *F*_(1,52.15)_ = 1.67, *p* = .20). There was no interaction of group by gain level for mean force (group × gain interaction: *F*_(2,104.37)_ = 0.10, *p* = .91). Mean force was greater in older participants (age main effect: *F*_(1,52.14)_ = 11.14, *p* < .005), though age-associated increases in mean force were similar across groups (group × age interaction: *F*_(1,51.99)_ = 0.61, *p* = .44).

**Fig 2.**
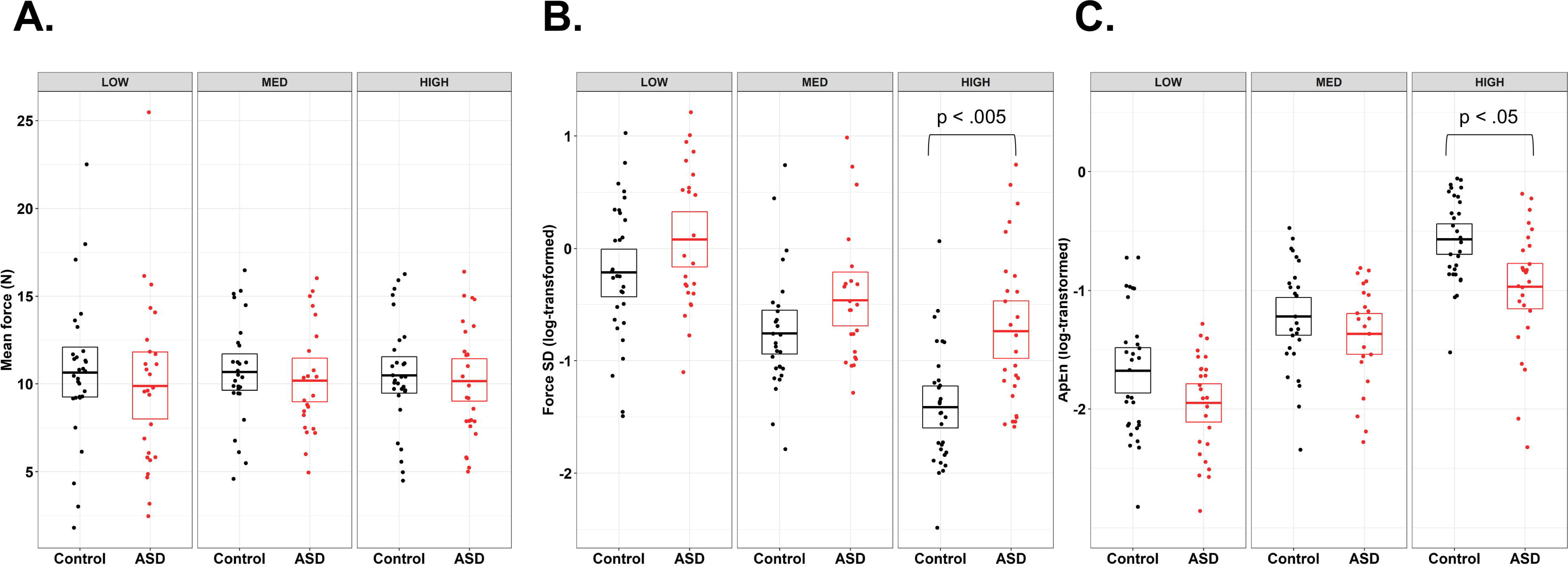
Results of the mixed effects models for behavioral force measures, controlling for age, sex, and IQ. A) Mean force was not different in individuals with ASD and TD controls, nor did it scale with gain. B) Force standard deviation (SD) significantly decreased with increasing gain (gain main effect), and was higher in individuals with ASD overall (group main effect). There was also a significant interaction of gain and group, such that individuals with ASD showed greater force SD compared to TD controls at high gain, but not at low or medium gain. C) Approximate Entropy (ApEn) significantly increased with increasing gain (gain main effect), and also was reduced in ASD relative to TD controls (group main effect). No gain by group interaction was observed, though post-hoc group comparisons are presented for ease of comparison.

Force SD decreased with increases in visual gain (Figure 2B; gain main effect: *F*_(2,103.82)_ = 80.10, *p* < .001). Individuals with ASD showed elevated force SD compared to TD controls (group main effect: *F*_(1,52.60)_ = 6.99, *p* = .01; Figure 2B), and group differences varied as a function of gain (group × gain interaction: *F*_(2,103.90)_ = 3.39, *p* = .04). Post-hoc analyses revealed that individuals with ASD showed increased force variability compared to TD controls at high gain (t_98.3_= 3.70, p < .005), but not at low (t_102.6_= 1.41, p = .72) nor medium gain (t_106.7_= 1.40, p = .73). Force SD decreased as a function of increased age (age main effect: *F*_(1,52.54)_ = 5.70, *p* = .02). Age-associated differences were similar across groups (group × age interaction: *F*_(1,52.33)_ = 0.13, *p* = .72).

ApEn increased with increases in visual gain (Figure 2C; gain main effect: *F*_(2,104.66)_ = 129.16, *p* < .001). Individuals with ASD showed reduced ApEn (i.e., reduced force entropy) compared to TD controls (Figure 2C; group main effect: *F*_(1,51.75)_ = 8.33, *p* < .01), and the magnitude of this difference was similar across gain levels (group × gain interaction: *F*_(2,104.75)_ = 1.45, *p* = .24). Increased age was associated with greater ApEn (age main effect: *F*_(1,51.87)_ = 9.53, *p* < .005), and age-associated increases in ApEn were similar across groups (group × age interaction: *F*_(1,51.60)_ = 1.05, *p* = .31).

### 3.2 Brain Activation Results

In the visuomotor – rest contrast, BOLD activation of multiple regions of the visuomotor network scaled with increases in visual gain, including bilateral posterior parietal cortex (V5, SPL), bilateral primary (M1) and premotor cortex, bilateral cerebellar Crus I, bilateral middle cingulate cortex, left middle occipital gyrus, and right supramarginal gyrus. Supplementary Figure 2 and Supplementary Table 2 show brain regions with BOLD activation that scaled with visual gain during visuomotor behavior (gain main effect).

Five regions showed significant group × gain level effects (Figure 3; Table 2), including SMA, bilateral SPL, left middle frontal gyrus (MFG), and left inferior frontal gyrus (IFG). Except for left IFG, brain activation in these regions scaled with gain level more strongly in individuals with ASD relative to TD controls resulting in increased activation in ASD compared to TD at high gain (Supplementary Table 3). For left IFG, this pattern of activation was similar, although the groups were not significantly diffrent at high gain. Additionally, there was a significant group × age effect in visual cortex (*F*_(1,50)_ = 15.30, *p* < .005). Activation in V1 increased with age in ASD, but decreased with age in TD controls.

**Fig 3.**
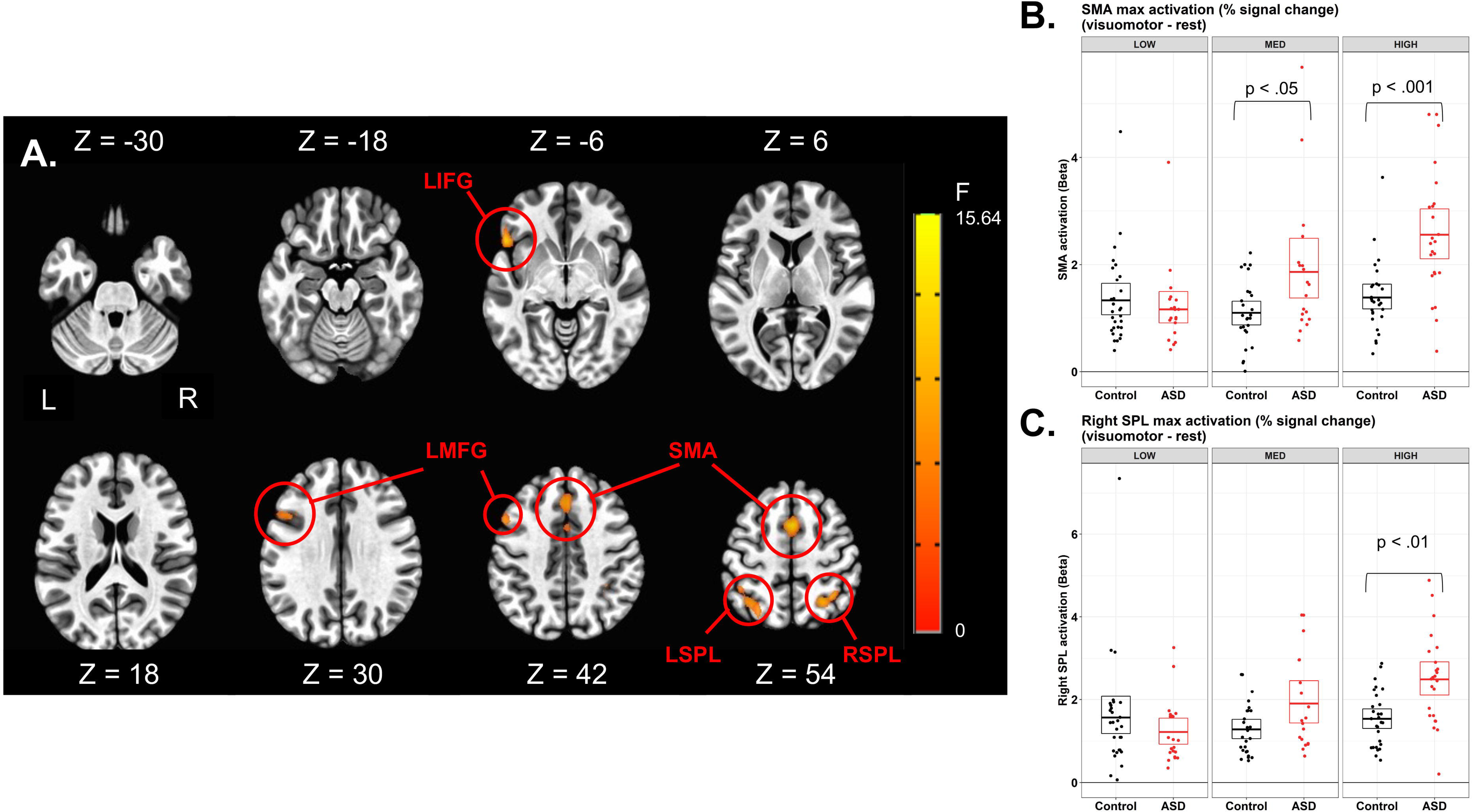
A) Axial slices showing regions with activation that scaled with gain level for visuomotor – rest contrasts for individuals with ASD more than for TD controls. The color bar ranges from F=0 to F=15.64, with an activation threshold of α < 0.05, corrected for multiple comparisons. B) Individuals with ASD show increased SMA activation relative to TD that scaled in severity with increases in gain. C) Right superior parietal lobule activation was greater in ASD compared to TD, especially during high gain.

**Table 2.**
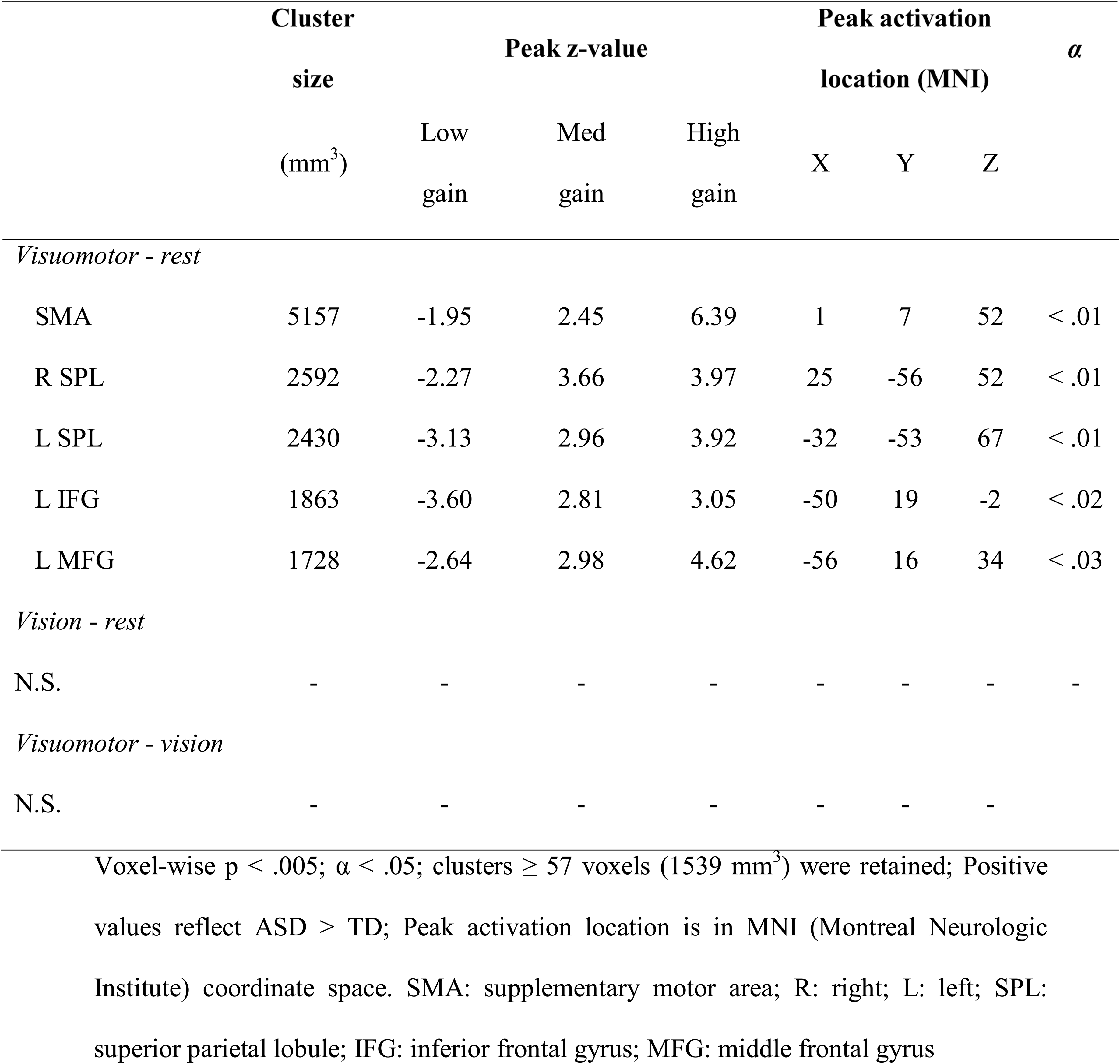
Brain regions showing significant group × visual gain interactions in activation in the linear mixed effects model (3dLME), controlling for age and sex

Brain activation results for visuomotor – vision and vision – rest contrasts are in Supplementary Results 1 and 2, respectively.

### 3.3 Visuomotor-Dependent Functional Connectivity Results

#### 3.3.1 Functional Connectivity Differences Between Individuals with ASD and TD Controls

Visuomotor-dependent functional connectivity between right IPL and a cluster in left prefrontal cortex extending from IFG into ventral premotor cortex (left PMv) differed between groups (Table 3; Figure 4). TD controls showed similar right IPL-left PMv connectivity during visuomotor action and rest, while individuals with ASD showed a visuomotor-dependent decrease in connectivity. Similarly, visuomotor-dependent functional connectivity between right IPL and left putamen was different between groups. TD controls showed similar right IPL-left putamen connectivity across visuomotor action and rest, while individuals with ASD showed reduced right IPL-left putamen connectivity during force compared to rest.

**Fig 4.**
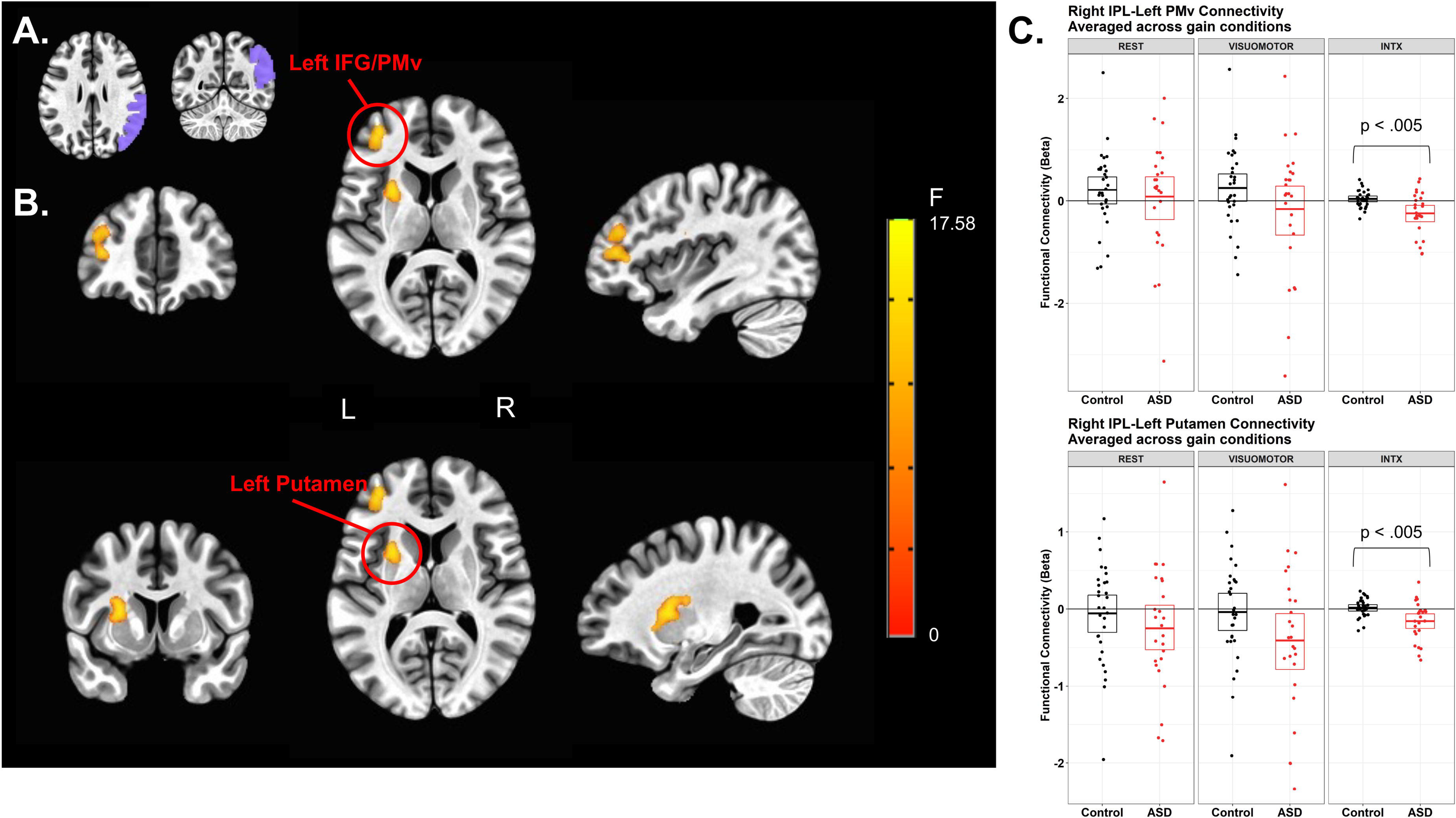
Results of the visuomotor-dependent connectivity analysis, corrected for age, sex, and IQ. A) The seed region used for the connectivity analysis – right inferior parietal cortex (IPL). B) Clusters in the left ventral premotor cortex and putamen showing significantly reduced visuomotor-dependent connectivity with right IPL in individuals with ASD compared to TD controls. The color bar ranges from F=0 to F=17.53, with an activation threshold of α < 0.05, corrected for multiple comparisons. C) The graphs display mean connectivity between the seed region and the cluster measured at rest and during visuomotor activity (VISUOMOTOR) for the two groups separately. The interaction (INTX) is the difference in connectivity for force minus rest, and is lower in ASD than TD controls in both clusters.

**Table 3.**
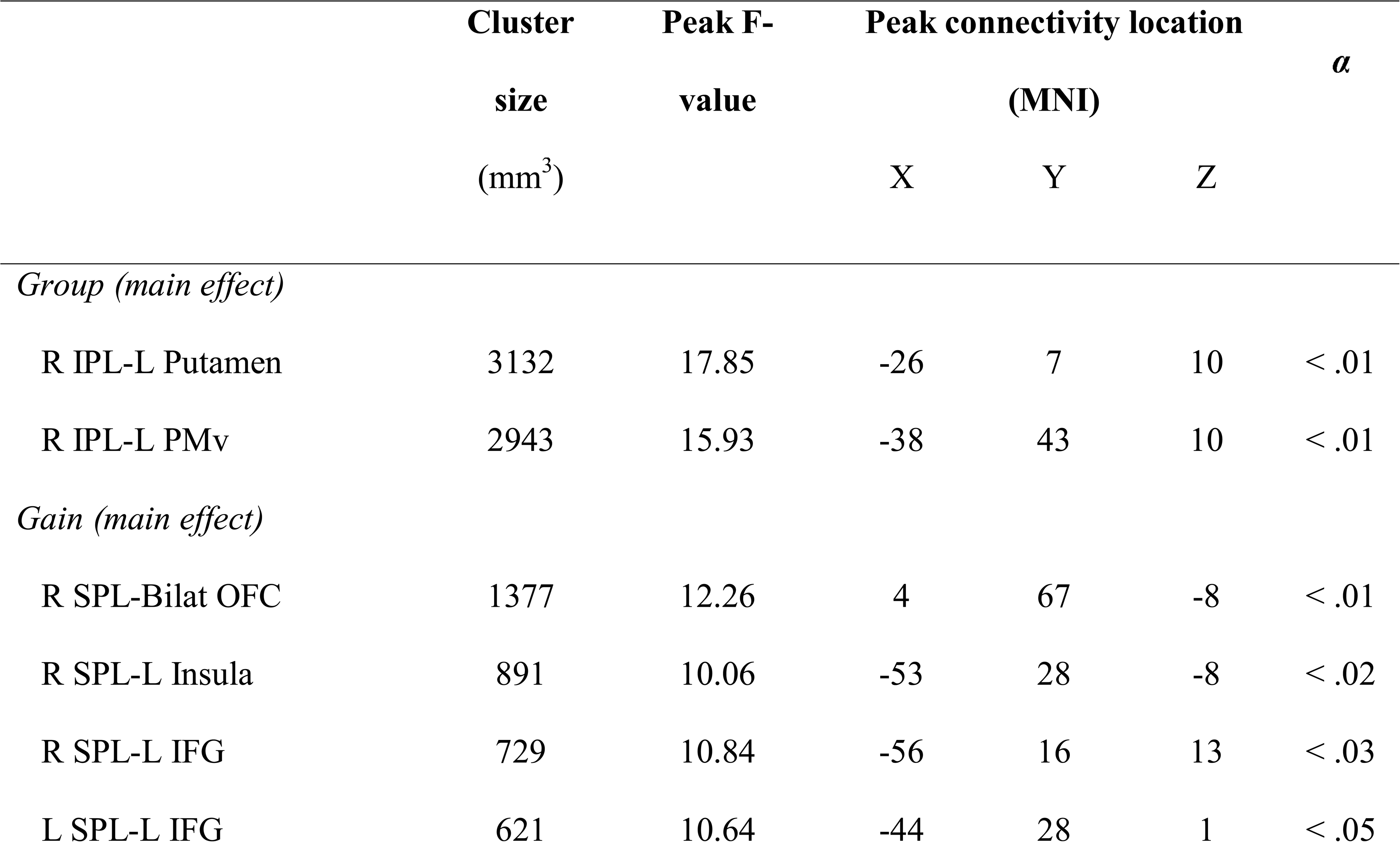
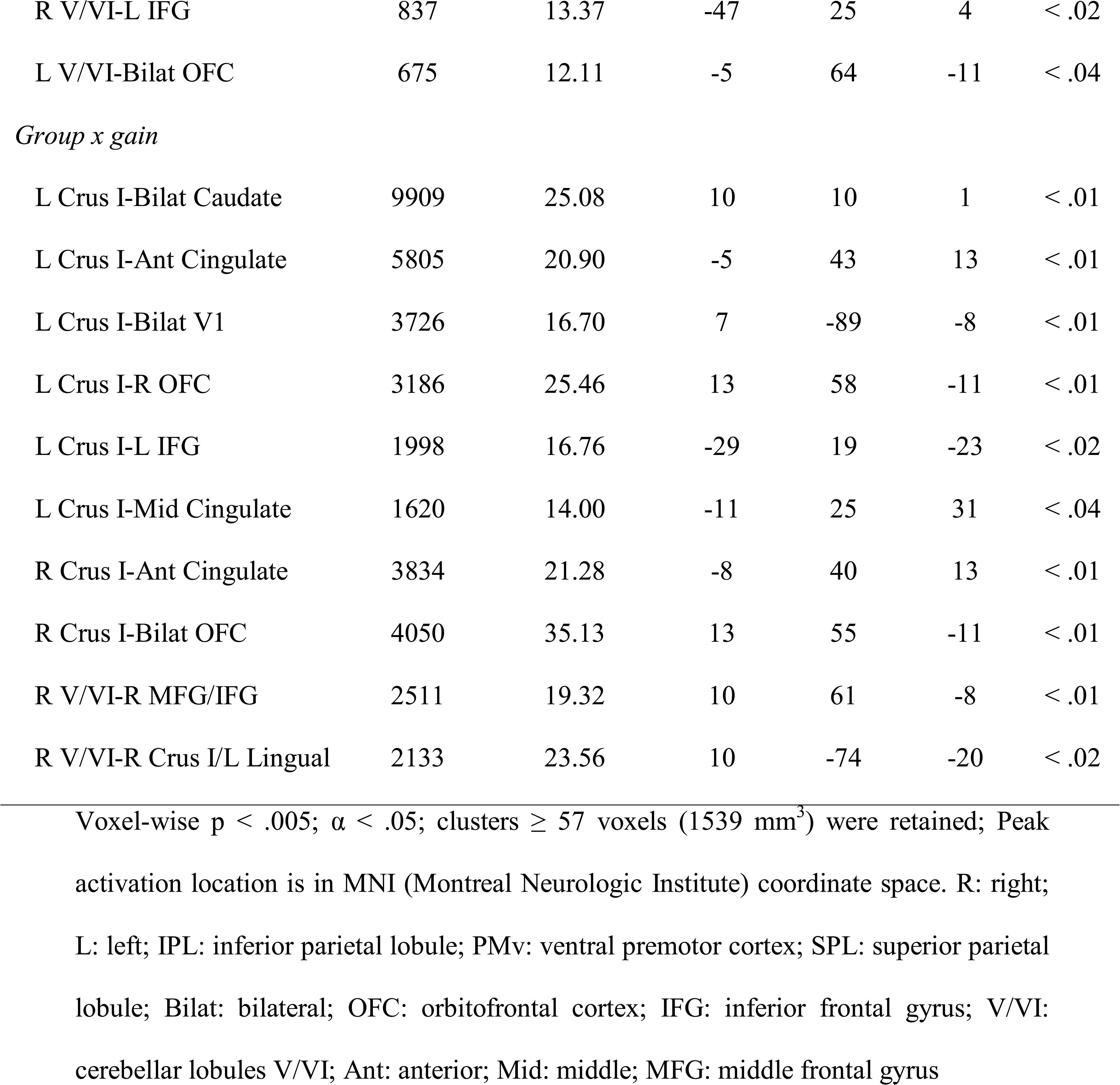
Brain regions showing significant group, gain, or group × age interactions in visuomotor-dependent functional connectivity in the linear mixed effects model (3dLME), controlling for age and sex

| | Cluster<br>size<br>(mm<sup>3</sup>) | Peak F-value | Peak connectivity location<br>(MNI) | | | $\alpha$ |
| --- | --- | --- | --- | --- | --- | --- |
|  |  |  | X | Y | Z |  |
| *Group (main effect)* |  |  |  |  |  |  |
| R IPL-L Putamen | 3132 | 17.85 | -26 | 7 | 10 | < .01 |
| R IPL-L PMv | 2943 | 15.93 | -38 | 43 | 10 | < .01 |
| *Gain (main effect)* |  |  |  |  |  |  |
| R SPL-Bilat OFC | 1377 | 12.26 | 4 | 67 | -8 | < .01 |
| R SPL-L Insula | 891 | 10.06 | -53 | 28 | -8 | < .02 |
| R SPL-L IFG | 729 | 10.84 | -56 | 16 | 13 | < .03 |
| L SPL-L IFG | 621 | 10.64 | -44 | 28 | 1 | < .05 |
| R V/VI-L IFG | 837 | 13.37 | -47 | 25 | 4 | < .02 |
| L V/VI-Bilat OFC | 675 | 12.11 | -5 | 64 | -11 | < .04 |
| *Group x gain* |  |  |  |  |  |  |
| L Crus I-Bilat Caudate | 9909 | 25.08 | 10 | 10 | 1 | < .01 |
| L Crus I-Ant Cingulate | 5805 | 20.90 | -5 | 43 | 13 | < .01 |
| L Crus I-Bilat V1 | 3726 | 16.70 | 7 | -89 | -8 | < .01 |
| L Crus I-R OFC | 3186 | 25.46 | 13 | 58 | -11 | < .01 |
| L Crus I-L IFG | 1998 | 16.76 | -29 | 19 | -23 | < .02 |
| L Crus I-Mid Cingulate | 1620 | 14.00 | -11 | 25 | 31 | < .04 |
| R Crus I-Ant Cingulate | 3834 | 21.28 | -8 | 40 | 13 | < .01 |
| R Crus I-Bilat OFC | 4050 | 35.13 | 13 | 55 | -11 | < .01 |
| R V/VI-R MFG/IFG | 2511 | 19.32 | 10 | 61 | -8 | < .01 |
| R V/VI-R Crus I/L Lingual | 2133 | 23.56 | 10 | -74 | -20 | < .02 |

Voxel-wise $p < .005$ ; $\alpha < .05$ ; clusters $\geq 57$ voxels (1539 mm<sup>3</sup>) were retained; Peak activation location is in MNI (Montreal Neurologic Institute) coordinate space. R: right; L: left; IPL: inferior parietal lobule; PMv: ventral premotor cortex; SPL: superior parietal lobule; Bilat: bilateral; OFC: orbitofrontal cortex; IFG: inferior frontal gyrus; V/VI: cerebellar lobules V/VI; Ant: anterior; Mid: middle; MFG: middle frontal gyrus

#### 3.3.2 Functional Connectivity Differences Across Visual Gain Levels

Visuomotor-dependent functional connectivity between right SPL and several frontal regions varied by gain, including bilateral OFC, left insula, and left IFG (Table 3). Visuomotor-dependent connectivity between right SPL and bilateral OFC as well as left insula was stronger during medium and high gain relative to low gain. Visuomotor-dependent connectivity between right SPL and left IFG was stronger during high relative to low gain. Similarly, visuomotor-dependent functional connectivity between left SPL and left IFG was stronger during high relative to low gain.

Visuomotor-dependent functional connectivity between right cerebellar lobules V/VI and left IFG varied by gain: visuomotor-dependent connectivity increased during high gain relative to low and medium gains (Table 3). Visuomotor-dependent functional connectivity between left cerebellar lobules V/VI and bilateral OFC was also greater during high and medium gain relative to low gain.

#### 3.3.3 Age-Associated Group Differences in Functional Connectivity

Left cerebellar Crus I and several regions showed significant group × age interactions in visuomotor-dependent functional connectivity, including bilateral caudate, anterior cingulate, bilateral V1, right OFC, left IFG, and middle cingulate. For bilateral caudate (*F*_(1,50)_ = 25.08, *p* < .001), anterior cingulate (*F*_(1,50)_ = 20.90, *p* < .001), and middle cingulate (*F*_(1,50)_ = 14.00, *p* < .005), connectivity with left Crus I during visuomotor action increased with age in individuals with ASD but remained stable across age in TD controls, resulting in increased visuomotor-dependent connectivity differences with age in ASD relative to TD controls. For left IFG (*F*_(1,50)_ = 16.76, *p* < .005), right OFC (*F*_(1,50)_ = 25.46, *p* < .001), and V1 (*F*_(1,50)_ = 16.70, *p* < .005), connectivity with left Crus I during visuomotor action increased with age in both groups but showed larger increases in individuals with ASD, resulting in increased visuomotor-dependent connectivity differences with age in ASD relative to TD controls (Figure 5A).

**Fig 5.**
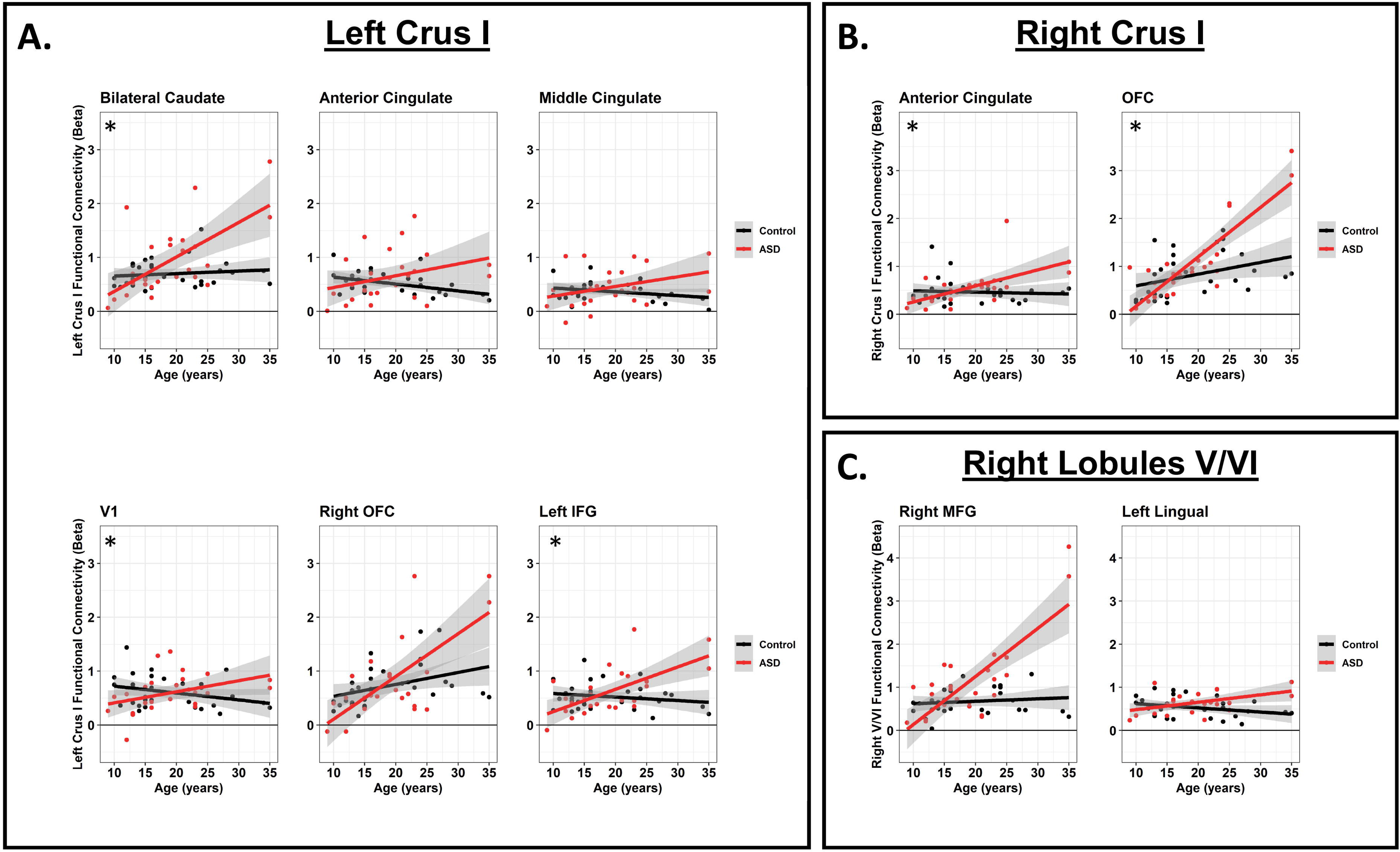
Age-related group differences in visuomotor-dependent connectivity. Scatterplots display the difference in connectivity between the two indicated brain regions during visuomotor activity versus rest (visuomotor – rest) for each participant. Red dots represent ASD participants, black dots represent TD control participants. Solid lines represent linear trend lines for age for each group, and shaded areas indicate 95% confidence intervals. Across most regions, ASD participants have reduced visuomotor-dependent connectivity at younger ages compared to TD control participants. Asterisks (*) in the upper-left portion of graphs denote group × age interactions which remained significant after excluding the four individuals in our data set > 30 years (2 ASD; 2 TD).

Right cerebellar Crus I and two prefrontal cortical regions showed significant group × age interactions in visuomotor-dependent functional connectivity, including anterior cingulate and bilateral OFC. For anterior cingulate (*F*_(1,50)_ = 21.28, *p* < .005) and OFC (*F*_(1,50)_ = 35.13, *p* < .001), connectivity with right Crus I increased with age more sharply in ASD than in TD controls (Figure 5B).

Right cerebellar lobules V/VI and two regions showed significant group × age interactions in visuomotor-dependent functional connectivity, including right middle and frontal gyri and a cluster spanning right medial cerebellum lobule V and left lingual gyrus. For right middle and frontal gyri (*F*_(1,50)_ = 19.32, *p* < .005), both groups showed increased connectivity with right lobules V/VI during visuomotor action with age, though this increase was larger in individuals with ASD. For right lobule V-left lingual gyrus (*F*_(1,50)_ = 23.56, *p* < .005), connectivity with right lobules V/VI during rest increased with age in individuals with ASD but showed larger increases with age in TD controls, while both groups showed similar increases in connectivity during visuomotor action with age. This resulted in increased visuomotor-dependent connectivity differences with age in ASD relative to TD controls (Figure 5C). Analyses excluding individuals > 30 years are in Supplementary Results 3, Supplementary Figure 4.

### 3.4 Relationships Between Visuomotor Behavior, Brain Function, and ASD Symptoms

There were no significant relationships between force SD and ApEn with ASD symptom severity, as measured by either the ADOS or the ADI (Supplementary Table 4).

Across participants and gain levels, increased left SPL activation during force was associated with reduced ApEn (*F*_(1,131.55)_ = 6.48, *p* = .01) and increased force SD (*F*_(1,135.54)_ = 14.08, *p* < .001). For individuals with ASD, reduced SMA activation during force was associated with increased severity of ADI-rated social (*F*_(1,11.87)_ = 10.83, *p* < .001) and communication abnormalities (*F*_(1,13.31)_ = 7.31, *p* = .02).

Increased visuomotor-dependent functional connectivity between left Crus I and bilateral V1 was associated with reduced force SD (*F*_(1,130.45)_ = 7.14, *p* < .01). Increased connectivity between right lobules V/VI and right MFG-IFG was associated with increased ApEn (*F*_(1,116.88)_ = 7.67, *p* < .01). The relationship between force SD and connectivity between right Crus I and anterior cingulate differed between groups (group × force SD; *F*_(1,130.93)_ = 11.94, *p* < .001); increased connectivity was associated with reduced force SD in TD controls but not in individuals with ASD. Increased ADOS-rated ASD symptom severity was associated with reduced connectivity between left Crus I and left IFG (*F*_(1,22)_ = 7.92, *p* = .01), and reduced connectivity between right lobules V/VI and right MFG-IFG (*F*_(1,22)_ = 7.15, *p* = .01).

## 4. DISCUSSION

We provide new evidence of atypical brain activation and functional connectivity associated with precision visuomotor impairments in ASD. Five key findings are highlighted. First, we show that increases in force SD are more severe at high visual gain suggesting impaired ability to process increased visual feedback. Second, we document that individuals with ASD show increased activation in SMA, bilateral SPL, and left MFG/IFG during visuomotor behavior. These differences were more pronounced at high visual gain suggesting reduced modulation of local circuit activity within sensory processing and motor control regions during precision behavior. Third, we report reduced visuomotor-dependent functional connectivity between right IPL and left PMv as well as right IPL and left putamen in ASD, implicating diminished integration of parietal feedback with cortical and subcortical motor planning systems. Fourth, we observe that individuals with ASD showed age-associated increases in functional connectivity of cerebellum and cerebral cortex, including occipital, medial prefrontal, and lateral prefrontal cortices, suggesting delayed cerebellar-cortical maturation in ASD. Last, we find that reduced cerebellar-prefrontal connectivity is associated with reduced force entropy and increased ASD symptoms, suggesting that atypical development of cerebellar-cortical networks may underpin sensorimotor and core social-communicative impairments.

### 4.1 Visuomotor Behavior in ASD

Consistent with our prior studies (Mosconi et al. 2015; Wang et al. 2015), we found that visually guided precision force variability is elevated in ASD, especially when visual feedback is amplified, suggesting that reduced visuomotor precision in ASD involves difficulty processing increased visual feedback information. Individuals with ASD also showed reduced entropy compared to TD controls implicating deficits integrating feedforward and multiple sensory feedback processes operating on different time scales.

Our results implicating deficient sensory feedback processing in ASD are consistent with prior studies demonstrating that patients may show atypical biases towards select sensory inputs, such as proprioceptive feedback, rather than integrating feedback across multiple sensory modalities (Haswell et al. 2009; Izawa et al. 2012). Results from the present study add evidence that sensorimotor impairment in ASD involves a reduced ability to integrate multisensory feedback to precisely and dynamically modulate behavior.

### 4.2 Visuomotor Brain Function in ASD

Activation across multiple brain regions scaled with increases in visual gain during visuomotor action, consistent with prior studies suggesting that the ability to reactively adjust ongoing sensorimotor behavior is dependent on scaling of activation across widely distributed cortical and cerebellar neural circuits (Vaillancourt et al. 2006b), involving posterior parietal and temporo-parietal cortex, motor cortex (Glickstein and Stein 1991), cerebellum, thalamo-M1 circuits (Calhoun et al. 2001), and striatal nuclei (Bostan et al. 2018). Individuals with ASD demonstrated atypical scaling of visuomotor network activation across visual gains, consistent with previous brain imaging findings during oculomotor movement (Takarae et al. 2007). Our finding relating SPL activation with increased force variability and reduced entropy provides additional evidence that visuomotor impairments in ASD reflect deficits in cortical processing of sensory feedback information. Together, these findings indicate that local activations of motor planning (SMA and MFG) and spatial processing (SPL) circuits show greater reactivity to amplifications of visual feedback that are associated with less precise visuomotor behavior.

### 4.3 Visuomotor-Dependent Functional Connectivity in ASD

Functional connectivity between IPL and PMv is associated with visually guided grasping (Bonini et al. 2010) and aids the reconciliation of multisensory incongruencies during visuomotor control (Schnell et al. 2007; Igelstrom and Graziano 2017). Reductions in the functional connectivity of this pathway during visuomotor control compared to rest in ASD indicates a selective decoupling of multisensory parietal input and motor planning circuits during behavior. These results suggest local sensory feedback processing and motor control circuits may function more independently in ASD than in TD and rely on local circuit modulation as reflected by increased activation of SMA, MFG and SPL, consistent with prior imaging studies (Just et al. 2007; Kana et al. 2009; Chen et al. 2015; Lee et al. 2016).

Our finding of reduced functional connectivity between right IPL and putamen in ASD implicates abnormal integration of visual feedback and motor timing and amplitude mechanisms during sensory guided motor refinement (Prodoehl et al. 2008). Consistent with our findings, multiple studies have documented functional and structural abnormalities of putamen in ASD, including reduced functional connectivity with V1 during finger tapping (Villalobos et al. 2005) and reduced volume associated with increased restricted and repetitive behaviors (Estes et al. 2011; Nickl-Jockschat et al. 2012).

### 4.4 Age-Associated Increases in Cerebellar-Cortical Functional Connectivity in ASD

Visuomotor-dependent cerebellar-striatal and cerebellar-cortical functional connectivity showed stronger age-associated increases in ASD relative to TD controls, suggesting the development of cerebellar processes involved in the modulation of timing and action selection striatal and prefrontal circuits is delayed in ASD (Yim et al. 2011). These results are consistent with prior studies showing atypical timing of motor output in children with ASD (D’Cruz et al. 2009; Wang et al. 2015), and associations between striatal volumes and repetitive behaviors that implicate deficits in efficient selection and assembly of motor programs (Langen et al. 2014).

Also implicated are the refinement and precision of behavioral output. Cerebellar Crus I-IFG circuits are involved in reactively adjusting motor commands in response to sensory feedback error information. Findings that functional connectivity of cerebellum and both ACC and OFC are reduced in children with ASD suggests delayed development of networks involved in flexibly modifying force output during goal-directed behavior. consistent with studies from our lab (Mosconi et al. 2015; Wang et al. 2015) and others (Morimoto et al. 2018; Lidstone et al. 2020) showing increased motor variability and reduced precision motor accuracy in ASD, especially in younger children. These precision sensorimotor tests may also be more challenging to individuals with ASD than TD based on studies showing that lateral cerebellum and prefrontal cortical circuits, including ACC and OFC, show greater coactivation during more challenging task or cognitive conditions (Braver et al. 1997; Rypma et al. 1999; Stoodley and Schmahmann 2009).

Visual cerebellar-cortical functional connectivity maturation also appears delayed in ASD. Visual input to cerebellum via pontine nuclei is used to guide adjustments of the motor command relayed to motor cortex (Glickstein 2000). Atypical development of this network implicates abnormal processing of visual feedback information that may contribute to increased variability and reduced entropy of sustained motor actions. Consistent with these findings, reduced cerebellar-cortical functional connectivity during rest (Ramos et al. 2018; Wang et al. 2019) and motor action (Mostofsky et al. 2009) have been reported in individuals with ASD, though increased cerebellar-cortical functional connectivity during rest also has been reported (Khan et al. 2015).

### 4.5 Visuomotor Processes, ASD Symptoms, and Motor Behavior

Cerebellar-prefrontal brain alterations in circuits supporting visuomotor behavior and higher-order language and socioemotional processing were related to visuomotor impairments and more severe core symptoms in ASD (Stoodley and Schmahmann 2009). The relationship of cerebellar-prefrontal functional connectivity to reduced force entropy and more severe social-communication abnormalities suggests that neurodevelopmental disruptions of these circuits may impact both early maturing sensorimotor behaviors and more complex social-communication abilities.

Correlations with motor behavior revealed that age-associated increases in cerebellar-prefrontal connectivity were related to better visuomotor control indicating reorganization of these network functions may compensate for deficits in visuomotor network connectivity and local circuit modulation. Cerebellar-anterior cingulate connectivity was associated with reduced force variability in TD controls but not in individuals with ASD. Relatedly, cerebellar-anterior cingulate connectivity increased with age in individuals with ASD. Anterior cingulate facilitates movement onset (Paus 2001) and prior studies have identified increased co-activation of anterior cingulate and cerebellum during finger tapping (Lench et al. 2017). These findings together suggest that, with age, individuals with ASD may selectively shift neural resources to cerebellar-anterior cingulate circuits during visuomotor behavior but fail to effectively utilize the circuit to improve movement precision.

### 4.6 Limitations

The present results should be considered in the context of several limitations. While task-based fMRI provides important advantages for tracking brain-behavior relationships relative to resting-state fMRI (Greene et al. 2018), this method presents limitations for assessing younger and more severely impaired individuals, including those with comorbid intellectual or developmental disabilities. Our results implicate distinct circuits that should be examined at younger ages and across the range of severity. Prior reports of increased sensorimotor impairments in individuals with comorbid ASD and intellectual or developmental disability relative to those with ASD alone suggest these circuits may be more aberrant in this population (Green et al. 2009). Our finding of age-dependent group differences in both task activation and functional connectivity highlights the need for longitudinal testing across early childhood into adulthood to determine primary and compensatory developmental mechanisms of sensorimotor and core deficits of ASD. Extended testing of middle-aged and older adults with ASD also will clarify the extent to which cerebellar-cortical networks mature across the lifespan.

### 4.7 Conclusions

We show that atypical sensorimotor behavior in ASD is linked to parietal, cerebellar and frontal dysfunction and varies according to the quality of sensory feedback. Combined with our findings that brain alterations are associated with visuomotor impairment and core ASD symptoms, our results suggest that reduced modulation of parietal-motor processing of sensory feedback information may contribute to multiple developmental disruptions in affected individuals.

## Data Availability

Data are available upon request from the corresponding author, Dr. Matthew Mosconi.

## Conflicts of Interest

Drs. Stephen Coombes and David Vaillancourt are co-founders and managers of Neuroimaging Solutions, LLC. The other authors have no competing interests to declare.

## Funding

This work was supported by the National Institutes of Health (grant numbers P50 ACE HD055751 (to J.A.S.), R01 MH112734 (to M.M.), K23 MH092696 (to M.M.), R21 AG065621 (to Z.W.), University of Florida CTSI Pilot Award UL1TR001427 (to Z.W.), U54 HD090216; and a Kansas Center for Autism Research and Training (K-CART) Research Investment Council Strategic Initiative Grant (to M.M.).

## Author Contributions

**Rebecca J. Lepping:** Data curation; Formal analysis; Writing - original draft. **Walker S. McKinney:** Data curation; Formal analysis; Writing - original draft. **Grant C. Magnon:** Writing - review & editing. **Sarah K. Keedy:** Investigation; Writing - review & editing. **Zheng Wang:** Data curation; Investigation; Software; Writing - review & editing. **Stephen A. Coombes:** Methodology; Software; Writing - review & editing. **David E. Vaillancourt:** Conceptualization; Methodology; Software; Writing - review & editing. **John A. Sweeney:** Conceptualization; Methodology; Funding acquisition; Writing - review & editing. **Matthew W. Mosconi:** Conceptualization; Funding acquisition; Supervision; Writing - review & editing.

We gratefully acknowledge the families who participated in these studies.

## Notes

### Author Declarations

Study procedures were reviewed and approved by the Institutional Review Board of the University of Illinois Chicago and abided by the Code of Ethics of the World Medical Association (Declaration of Helsinki).

