## Supplementary Material for "Visuomotor brain network activation and functional connectivity among individuals with autism spectrum disorder"

Table of Contents

1. Supplementary Results 1. Brain activation for vision – rest contrasts.
2. Supplementary Results 2. Brain activation for visuomotor – vision contrasts.
3. Supplementary Results 3. Age-Associated Group Differences in Functional Connectivity Excluding Individuals > 30 Years
4. Supplementary Figure 1. Regions of the sensorimotor integration network interrogated as seed regions for context-dependent functional connectivity psychophysiological interaction (PPI) analysis: Bilateral Inferior and Superior Parietal Cortices, Cerebellar Lobules V/VI and Crus I.
5. Supplementary Figure 2. Brain activation that scaled with visual feedback gain during visuomotor – rest contrasts. Activation in these regions significantly increased with increases in gain.
6. Supplementary Figure 3. Brain activation that scaled with visual feedback gain during vision – rest contrasts. Activation in these regions significantly increased with increases in gain.
7. Supplementary Figure 4. Age-related group differences in visuomotor-dependent connectivity which remained significant after excluding the four individuals in our data set > 30 years (2 ASD; 2 TD).
8. Supplementary Table 1. Mean motion censoring of fMRI data by gain level for ASD and TD controls.
9. Supplementary Table 2. Brain regions showing activation that scaled with visual feedback gain during visuomotor – rest, vision – rest, and visuomotor – vision contrasts.
10. Supplementary Table 3. Post-hoc group pairwise comparisons of brain activation in regions showing significant group x gain interactions in the whole brain 3dLME model of visuomotor – rest.
11. Supplementary Table 4. Associations between sensorimotor behavior and clinical outcomes.
12. Supplementary Table 5. Associations between sensorimotor behavior and brain activation in regions showing significant group x gain interactions in the whole brain 3dLME model of visuomotor – rest.
1. **Supplementary Results 1. Brain Activation for Vision – Rest Contrasts.** For vision – rest contrasts, BOLD activation for multiple brain regions scaled with gain level, including bilateral V5, right SPL, left dorsolateral prefrontal cortex (DLPFC), left supramarginal gyrus, left lingual gyrus, and right ventral premotor cortex (Supplementary Figure 3, Supplementary Table 2). No brain regions showed differences in BOLD activation between groups (ASD vs. TD), and no regions showed significant group × gain level effects.
2. **Supplementary Results 2. Brain Activation for Visuomotor – Vision Contrasts.** Activation in multiple regions of the visuomotor network scaled with visual gain after controlling for activation specific to visual feedback (Supplementary Table 2). SMA, left M1, and left SMG all showed increased activation with increases in visual gain. Right S1 showed the opposite pattern, with activation decreasing as gain level was increased. No brain regions showed differences in BOLD activation between groups (ASD vs. TD) or group × visual gain level interactions. One cluster in the anterior cingulate cortex showed a significant group × age interaction, with TD participants showing a negative correlation with age, and ASD participants showing no relationship between age and activation.
3. **Supplementary Results 3. Age-Associated Group Differences in Functional Connectivity Excluding Individuals > 30 Years.** Based on the skewed age distribution of our sample, we repeated age analyses excluding the four individuals in our data set > 30 years (2 ASD; 2 TD). These results were substantively similar but indicated that individuals with ASD showed reduced visuomotor-dependent functional connectivity at younger ages but similar levels of functional connectivity during adolescence and early adulthood. Five of the pathways tested also no longer showed significant group x age interactions: visuomotor-dependent connectivity between 1) left Crus I and anterior cingulate cortex, 2) left Crus I and middle cingulate cortex, 3) left Crus I and right OFC, 4) right lobules V/VI and right MFG, and 5) right lobules V/VI and left lingual gyrus (Supplementary Figure 4).
4. **Supplementary Figure 1. PPI seed regions.** Eight hypothesis-driven regions of interest (ROIs) were used as seed regions for visuomotor-dependent functional connectivity analyses. Cortical ROIs were obtained using Brainnetome (Fan et al., 2016). Cerebellar ROIs were obtained using the spatially unbiased template (SUIT) cerebellar atlas (Diedrichsen, 2006).
5. **Supplementary Figure 2. Visuomotor – Rest Gain Main Effect.** A) Axial slices showing scaling of blood oxygenation level dependent (BOLD) activation with gain level for visuomotor – rest contrasts for all participants. The color bar ranges from F=0 to F=44.91, with an activation threshold of α *<* 0.05, corrected for multiple comparisons. B) The graph shows activation in the superior parietal cortex (SPL) – a pattern that was exemplified across the significant regions with activation increasing with increases in gain.
6. **Supplementary Figure 3. Vision – Rest Gain Main Effect.** Axial slices showing scaling of blood oxygenation level dependent (BOLD) activation with gain level for vision – rest contrasts for all participants. The color bar ranges from F=0 to F=32.39, with an activation threshold of α *<* 0.05, corrected for multiple comparisons.
7. **Supplementary Figure 4.** **Age-related group differences in visuomotor-dependent connectivity which remained significant after excluding the four individuals in our data set > 30 years (2 ASD; 2 TD).** Scatterplots display the difference in connectivity between the two indicated brain regions during visuomotor activity versus rest (visuomotor – rest) for each participant. Red dots represent ASD participants, black dots represent TD control participants. Solid lines represent linear trend lines for age for each group, and shaded areas indicate 95% confidence intervals. Y axes are scaled relative to each subpanel.
8. **Supplementary Table 1. Mean motion censoring of fMRI data by gain level for ASD and TD controls.**

|  | Low gain | Medium gain | High gain |
| --- | --- | --- | --- |
| *Percent Volumes Censored* |  |  |  |
| ASD | 9.97% | 6.67% | 8.90% |
| TD control | 4.88% | 5.04% | 4.31% |
| *Average Motion Per Volume* |  |  |  |
| ASD | 0.10 mm | 0.08 mm | 0.09 mm |
| TD control | 0.08 mm | 0.08 mm | 0.08 mm |

9. **Supplementary Table 2. Regions of interest showing activation that scaled with visual feedback gain during visuomotor – rest, vision – rest, and visuomotor – vision contrasts**

|  | **Cluster size** | **Peak *F* value** | **Peak activation location** | | | ***α*** |
| --- | --- | --- | --- | --- | --- | --- |
|  | (mm^3^) |  | X | Y | Z |  |
| **Visuomotor – Rest** |  |  |  |  |  |  |
| R V5 | 19872 | 44.66 | 52 | -71 | 7 | < .01 |
| L V5 | 17361 | 35.04 | -50 | -74 | 7 | < .01 |
| L V1 | 1404 | 12.14 | -26 | -95 | 4 | < .01 |
| L Mid Occipital Gyrus | 702 | 14.19 | -26 | -86 | 34 | < .03 |
| R SPL | 6615 | 22.92 | 13 | -62 | 67 | < .01 |
| L SPL | 16146 | 25.99 | -38 | -50 | 58 | < .01 |
| Bilateral Crus I | 2808 | 12.97 | 1 | -86 | -29 | < .01 |
| R M1 | 18252 | 30.63 | 43 | -8 | 55 | < .01 |
| L M1 | 7344 | 36.08 | -44 | -8 | 55 | < .01 |
| R PMv | 15417 | 31.84 | 61 | 10 | 25 | < .01 |
| L PMv | 13743 | 23.60 | -59 | 10 | 28 | < .01 |
| R SMG | 6966 | 20.88 | 55 | -32 | 25 | < .01 |
| L Mid Cingulate | 1431 | 23.90 | -14 | -26 | 43 | < .01 |
| R Mid Cingulate | 891 | 15.47 | 16 | -26 | 37 | < .02 |
| **Vision – Rest** |  |  |  |  |  |  |
| R V5 | 15471 | 32.39 | 49 | -68 | 1 | < .01 |
| L V5 | 7992 | 19.27 | -53 | -74 | 1 | < .01 |
| R SPL | 3105 | 13.08 | 31 | -41 | 70 | < .01 |
| R PMv | 675 | 11.21 | 55 | 10 | 22 | < .04 |
| L DLPFC | 2160 | 13.58 | -47 | 46 | 7 | < .01 |
| L SMG | 1944 | 14.74 | -65 | -23 | 34 | < .01 |
| L Lingual Gyrus | 945 | 11.08 | -11 | -41 | -2 | < .01 |
| **Visuomotor – Vision** | |  |  |  |  |  |
| SMA | 2430 | 17.73 | -2 | -14 | 61 | < .01 |
| L M1 | 1404 | 16.11 | -44 | -11 | 58 | < .01 |
| R S1 | 648 | 15.02 | 34 | -29 | 64 | < .01 |
| L SMG | 621 | 11.33 | -44 | -35 | 22 | < .05 |

voxel-wise p < .001; α < .05; clusters ≥ 23 voxels

R: right; L: left; V5: primary visual cortex; M1: precentral gyrus; SPL: superior parietal lobule; PMv: ventral premotor cortex; SMG: supramarginal gyrus; Mid: middle; OFC: orbitofrontal cortex; DLPFC: dorsolateral prefrontal cortex; SMA: supplementary motor area; S1: postcentral gyrus; MFG: middle frontal gyrus

10. **Supplementary Table 3. Post-hoc group pairwise comparisons of maximum brain activation in regions identified as showing significant group × gain interactions in the whole brain 3dLME model of visuomotor – rest.**

| **Visuomotor – Rest** |  |  |  |  |
| --- | --- | --- | --- | --- |
| **SMA** | **Gain** | **Estimate (SE)** | ***t* ratio *(df)*** | ***p*** |
|  | Low | -0.15 (0.25) | -0.58 (131.8) | .99 |
|  | Medium* | 0.80 (0.27) | 3.01 (135.0) | < .05 |
|  | High* | 1.18 (0.25) | 4.77 (130.7) | < .001 |
| **RSPL** | Low | -0.30 (0.28) | -1.08 (128.7) | .89 |
|  | Medium | 0.67 (0.29) | 2.30 (132.8) | .20 |
|  | High* | 0.96 (0.27) | 3.58 (127.2) | < .01 |
| **LSPL** | Low | -0.22 (0.25) | -0.90 (127.0) | .95 |
|  | Medium | 0.59 (0.26) | 2.26 (131.4) | .22 |
|  | High* | 0.84 (0.24) | 3.44 (125.3) | < .05 |
| **LMFG** | Low | -0.13 (0.17) | -0.77 (117.4) | .97 |
|  | Medium | 0.42 (0.18) | 2.42 (123.7) | .16 |
|  | High* | 0.57 (0.16) | 3.49 (114.7) | < .01 |
| **LIFG** | Low | -0.33 (0.28) | -1.16 (117.6) | .86 |
|  | Medium | 0.42 (0.30) | 1.42 (123.8) | .72 |
|  | High | 0.69 (0.28) | 2.52 (114.9) | .13 |

*Tukey-adjusted *p* < .05; Estimates reflect percent signal change of designated contrast; Positive Estimates reflect ASD > TD controls.

11. **Supplementary Table 4. Associations between sensorimotor behavior and clinical outcomes.**

|  | **Clinical Outcome** | ***ρ*** | ***F (DF,DF)*** | ***p*** |
| --- | --- | --- | --- | --- |
| **Force SD** | ADOS CSS | .04 | 0.13 (1,23.76) | .72 |
|  | ADI-R (A) | -.24 | 1.18 (1,14.69) | .30 |
|  | ADI-R (Verbal B) | -.11 | 0.43 (1,14.60) | .52 |
|  | ADI-R (C) | -.08 | 0.02 (1,16.39) | .89 |
| **ApEn** | ADOS CSS | -.09 | 0.10 (1,22.94) | .75 |
|  | ADI-R (A) | .04 | 0.18 (1,14.23) | .68 |
|  | ADI-R (Verbal B) | -.01 | 0.06 (1,14.08) | .81 |
|  | ADI-R (C) | -.15 | 0.17 (1,14.09) | .69 |

Significance was determined from using separate linear mixed effects models for each clinical outcome which included fixed effects for gain and a gain × clinical outcome interaction term, although Spearman rank-order correlation coefficients (calculated by averaging sensorimotor outcomes across gain levels) are reported to aid in directional interpretation. Non-significant gain × clinical outcome interaction terms were removed to maintain model parsimony but are reported when significant. ADOS: Autism Diagnostic Observation Schedule; CSS: Calculated severity score; ADI-R; Autism Diagnostic Interview-Revised; * *p* < .05.

12. **Supplementary Table 5. Associations between sensorimotor behavior and brain activation in regions showing significant group × gain interactions in the whole brain 3dLME model of visuomotor – rest.**

|  | **Brain Region** | ***r*** | ***F (DF,DF)*** | ***p*** |
| --- | --- | --- | --- | --- |
| **Force SD** | SMA | .30 | 2.00 (1,137.53) | .16 |
|  | R SPL | .23 | 0.16 (1,137.96) | .69 |
|  | L SPL* | .34 | 14.08 (1,135.54) | < .001 |
|  | L MFG | .21 | 0.01 (1,135.43) | .92 |
|  | L IFG | .17 | 0.25 (1,134.80) | .61 |
| **ApEn** | SMA | -.21 | 0.04 (1,128.25) | .84 |
|  | R SPL | -.25 | 1.14 (1,131.04) | .29 |
|  | L SPL* | -.23 | 6.48 (1,131.55) | .01 |
|  | L MFG | -.17 | 0.15 (1,138.65) | .70 |
|  | L IFG | -.19 | 0.32 (1,138.84) | .57 |

Significance was determined from using separate linear mixed effects models for max activation within each brain region which included fixed effects for gain and a gain × activation interaction term, although Pearson correlation coefficients (calculated by averaging sensorimotor outcomes and activation across gain levels) are reported to aid in directional interpretation. Non-significant gain × activation interaction terms were removed to maintain model parsimony but are reported when significant. SMA: supplementary motor area; R: right; SPL: superior parietal lobule; L: left; MFG: middle frontal gyrus; IFG: inferior frontal gyrus; * *p* < .05.
