## Supplementary figures and images for "Visuomotor brain network activation and functional connectivity among individuals with autism spectrum disorder"

### Supplementary Figure 1

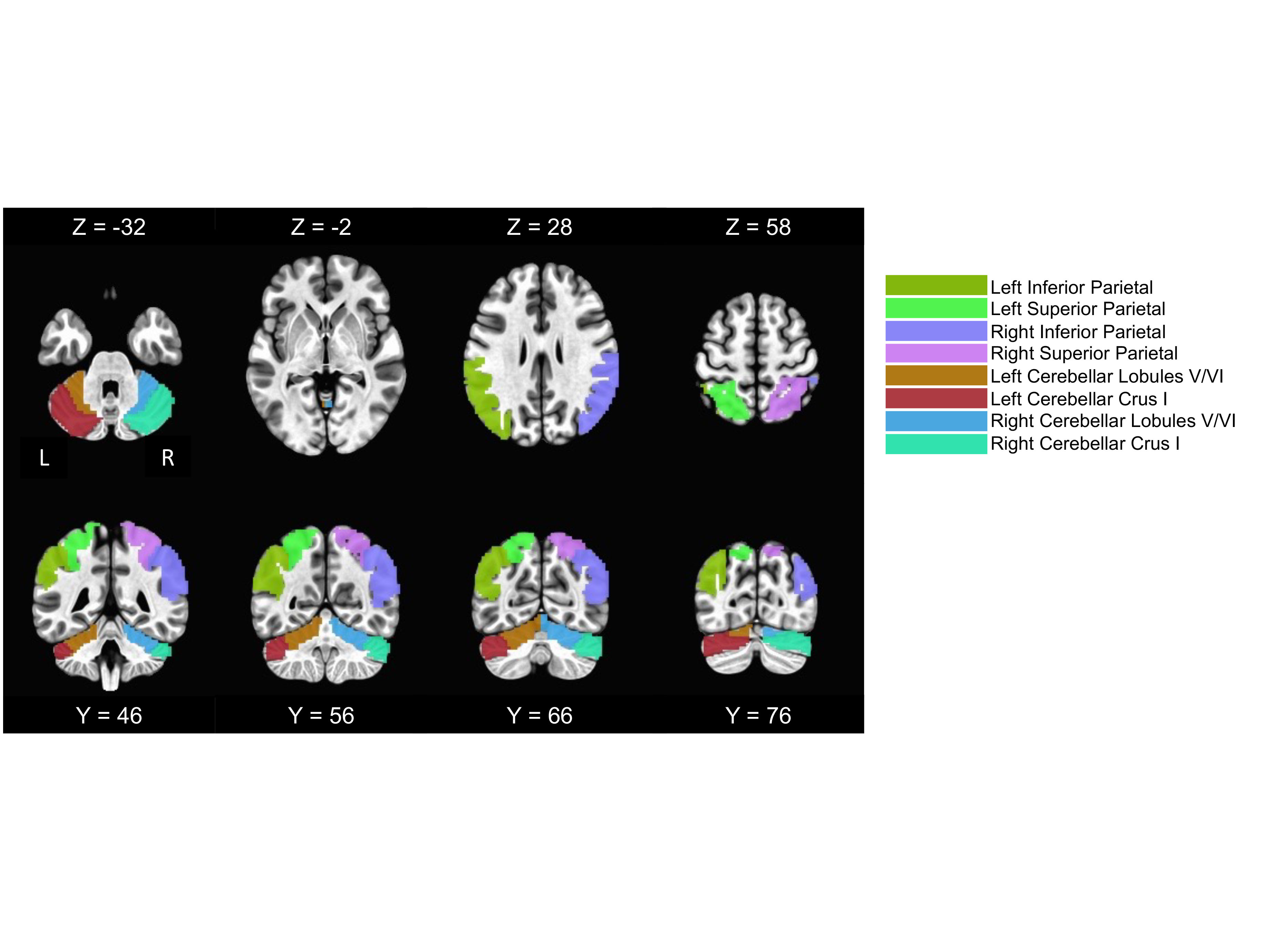

### Supplementary Figure 2

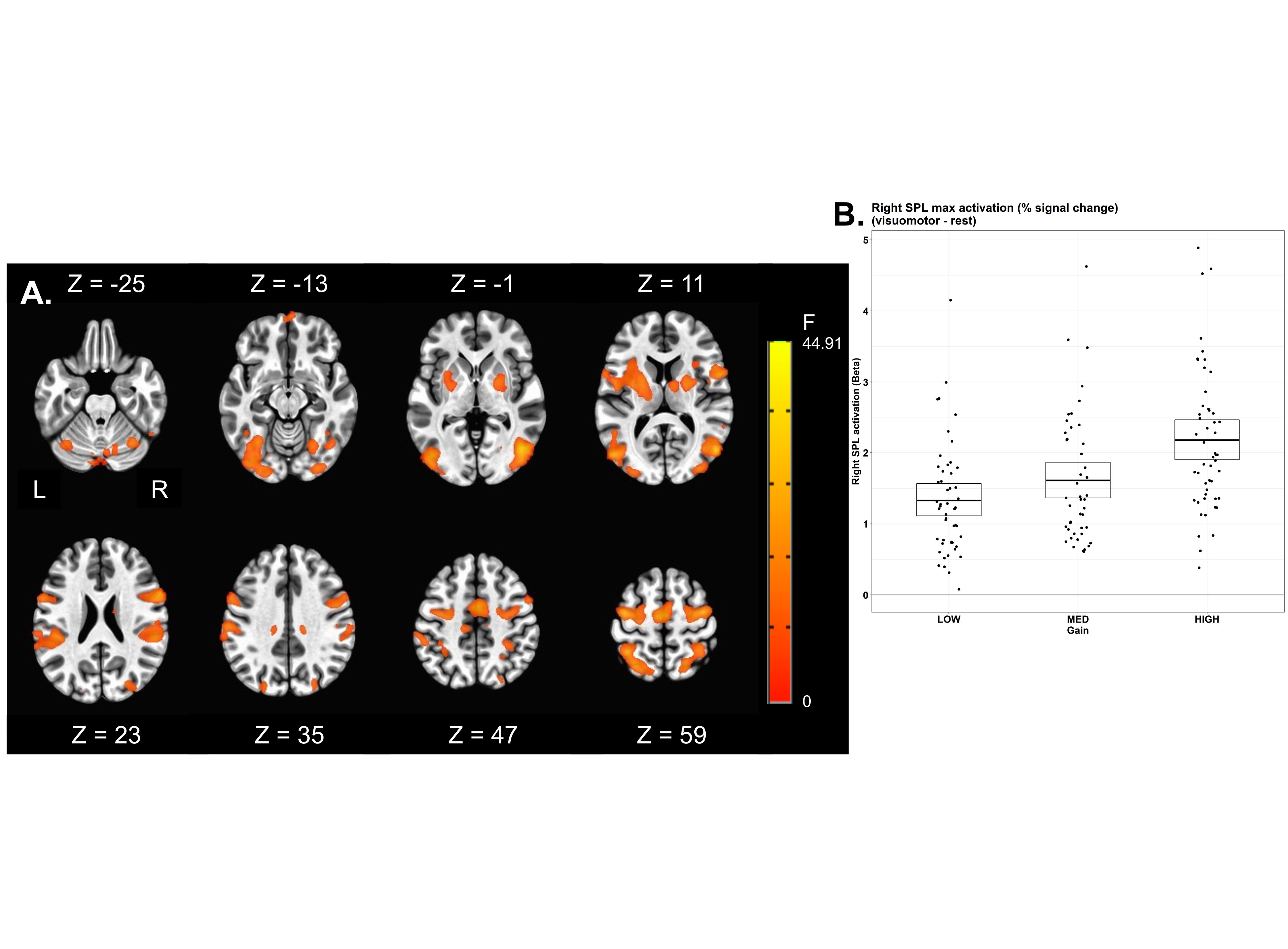

### Supplementary Figure 3

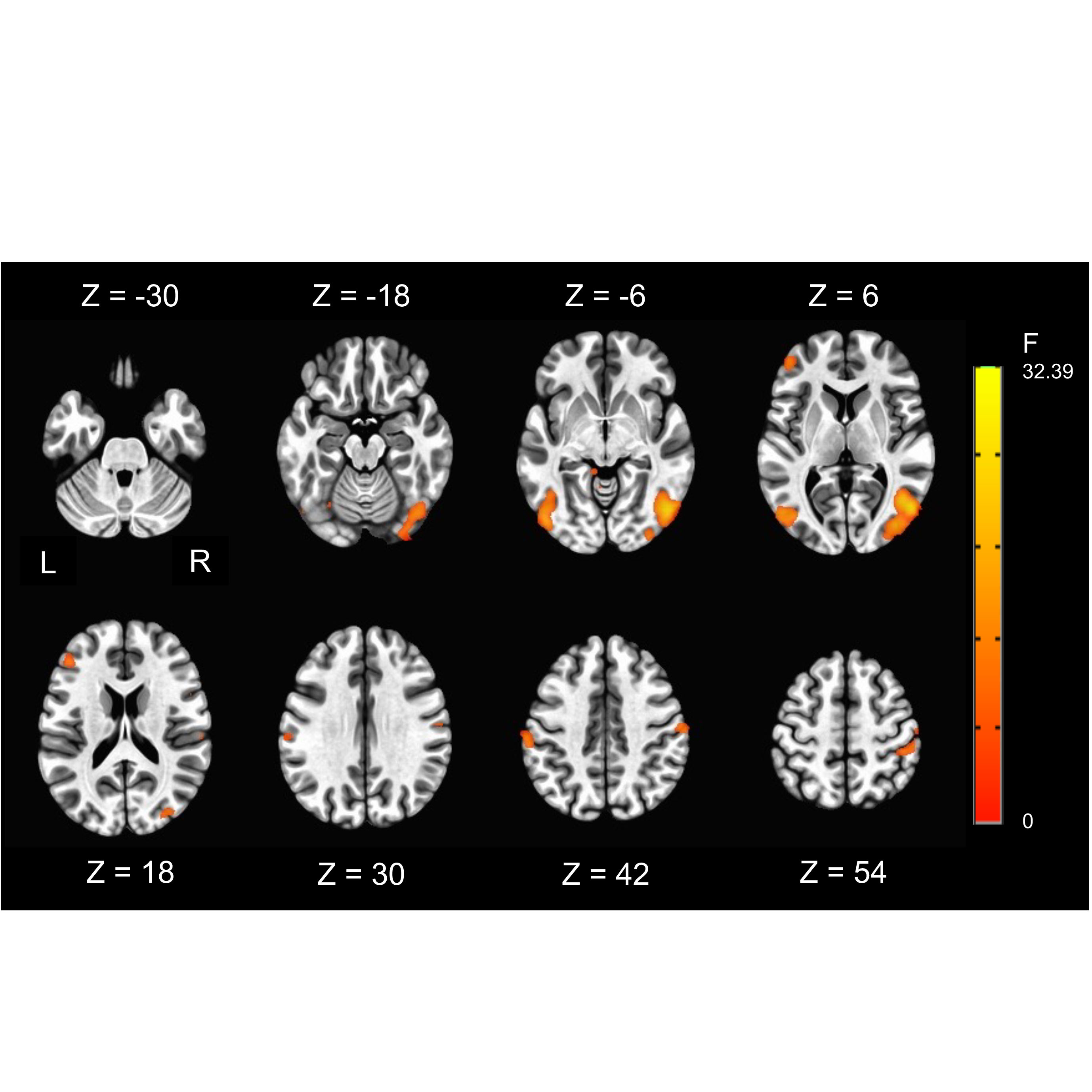

### Supplementary Figure 4

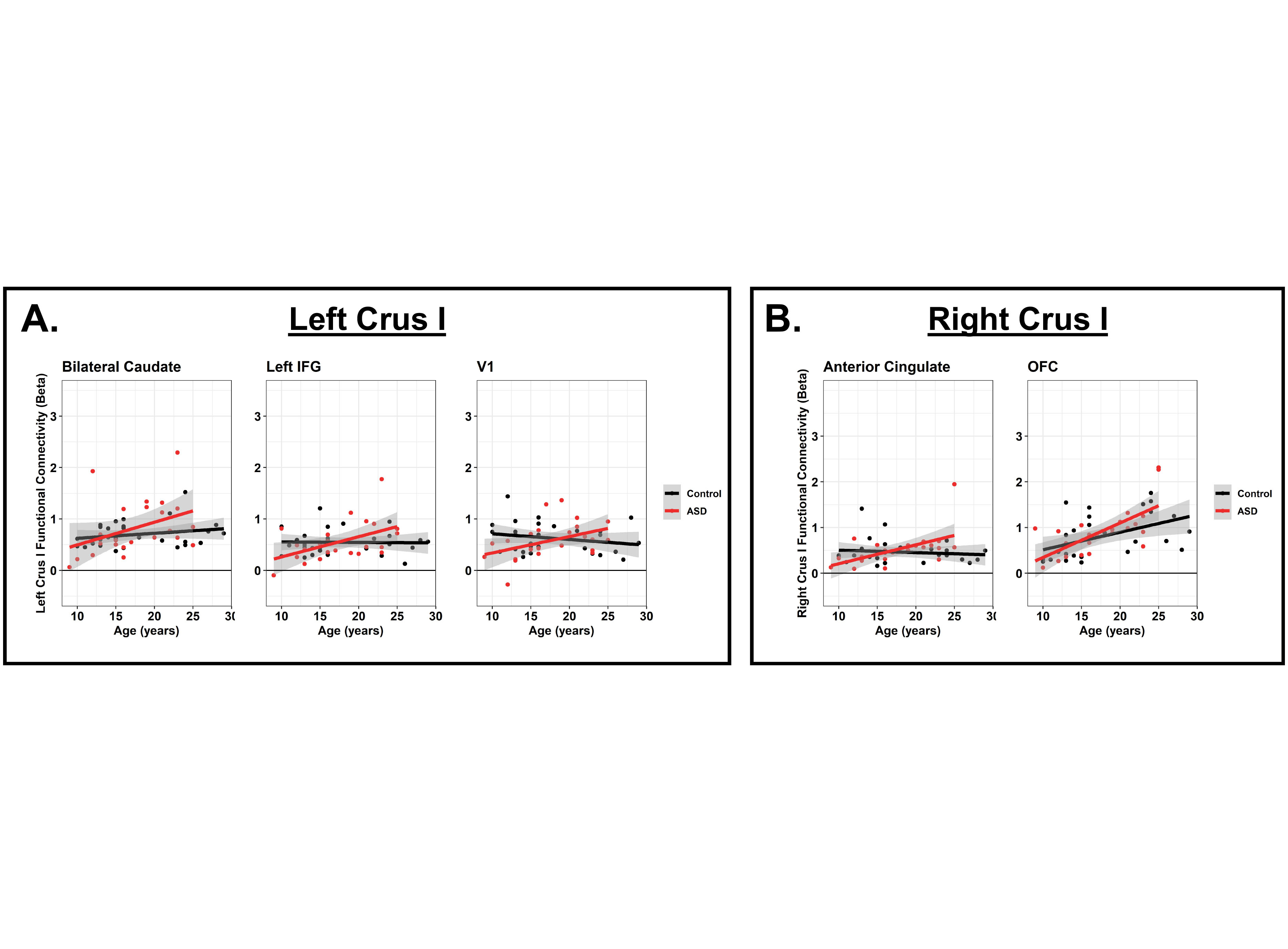
